# Treatment Outcome in Patients With Spinal Neurocysticercosis: A Systematic Review of Published Cases and Case Series

**DOI:** 10.1101/2024.07.24.24310906

**Authors:** Ravindra Kumar Garg, Imran Rizvi, Harish Nigam, Shweta Pandey, Ravi Uniyal

## Abstract

**Background:** Spinal neurocysticercosis is a rare central nervous system infection caused by the larval form of the *Taenia solium* tapeworm. Due to its rarity, most knowledge is derived from isolated case reports.

**Objectives:** This systematic review aims to evaluate existing case reports and observational studies to provide a comprehensive overview of the disease’s clinical presentation, and treatment outcomes.

**Methods:** Adhering to PRISMA guidelines, a search was conducted across multiple databases including PubMed, Scopus, Embase, and Google Scholar. Case reports, case series, and observational studies were included. The review is registered with PROSPERO (CRD42024496957).

**Results:** The search yielded 163 records describing 197 patients. Symptoms ranged from one week to over three years, with the most common being paraparesis or quadriparesis (61%) and back pain. Treatment modalities varied. with a combination of surgery and cysticidal drugs being the most preferred (45.2%) treatment. Surgery was done in 77% (152/197) of cases. In 45 % of cases (89/197) cysticidal drugs were given following surgery. Medical treatment alone was given to 22% (43/197) of patients.

The majority of cases (92%) irrespective of treatment modality showed clinical improvement. Post-operative complications caused three deaths.

**Conclusions:** We noted that surgery followed by cysticidal drugs was the most preferred treatment. Medical treatment alone was given to many patients. Clinical improvement was observed in most cases, regardless of the treatment option used. The use of cysticidal drugs could eliminate the need for surgery in many spinal neurocysticercosis patients.

**Key message:** Most spinal neurocysticercosis patients improve clinically with surgery and cysticidal drugs, the most common treatment. Medical management alone also benefits many, suggesting potential to reduce surgical intervention with effective drug therapy.

## Introduction

Neurocysticercosis, a preventable parasitic disease affecting the central nervous system, is caused by the pork tapeworm, *Taenia solium*. Human infection typically follows after the ingestion of the consumption of food or water contaminated with *Taenia solium* ova. It is believed that the total number of individuals affected by neurocysticercosis, encompassing both those showing symptoms and those without, lies somewhere between 2.56 and 8.30 million. This estimation is based on the varied prevalence data available for epilepsy.^1,2^

*Taenia solium* larvae may preferentially affect the muscles, skin, eyes, and the brain. Spinal neurocysticercosis is a rarer form of the disease.^2^ The exact proportion of spinal cord involvement among neurocysticercosis cases is not precisely known.

Barrie and colleagues cited a rate of spinal involvement ranging between 0.7% and 5.85% of all neurocysticercosis cases.^3^

Treatment modalities for spinal neurocysticercosis involve a combination of anti- parasitic medications like albendazole or praziquantel, corticosteroids to reduce inflammation and surgical intervention. Most of the information about spinal neurocysticercosis is available only in the form of isolated cases. We aimed to systematically review all reported cases to evaluate the efficacy of various treatment modalities.

## Methods

This systematic review was conducted following the PRISMA (Preferred Reporting Items for Systematic Reviews and Meta-Analyses) guidelines, as specified by the PRISMA checklist. The review has been recorded with PROSPERO, carrying the registration number CRD42024496957.^4^ There was no requirement for approval from the Institutional Ethics Committee for this review, as it did not involve human participants.

### Search strategy

Relevant reports were identified through comprehensive searches in the PubMed, Scopus, Embase, and Google Scholar databases. The first 50 pages of results on Google Scholar were meticulously examined for applicable studies. The search process was not confined by language, and for articles not in English, “A Google translator” was employed to translate them into English. Our search methodology was as follows: (((Spinal cord) OR (intramedullary)) OR (Extramedullary)) AND (Cysticercosis/ OR Neurocysticercosis). The searches were concluded on January 2, 2024.

### Eligibility criteria

The diagnostic criteria for neurocysticercosis are classified into four categories: absolute, major, minor, and epidemiological, which aid in the determination of definitive, probable, and possible diagnoses. A definitive diagnosis can be confirmed through one of the following methods: histological verification of the parasite from a biopsy sample, direct visual identification of a parasite in the eye, or computed tomography/magnetic resonance imaging that reveals cystic lesions containing the characteristic scolex of the parasite. Additionally, a definitive diagnosis may be made by a combination of two major criteria, or one major criterion along with two minor and one epidemiological criterion. A probable diagnosis is reached using one of these combinations: one major criterion and two minor criteria, one major criterion alongside one minor and one epidemiological criterion, or three minor criteria in conjunction with one epidemiological criterion. A possible diagnosis, which is less definitive, is made with either one major criterion, two minor criteria, or one minor and one epidemiological criterion. The major criteria include neuroimaging results indicative of neurocysticercosis, positive tests for anticysticercal antibodies, or X-ray evidence of characteristic calcifications in the muscles. The minor criteria encompass the presence of subcutaneous nodules calcifications seen on X-ray, clinical indications of neurocysticercosis, or resolution of lesions after treatment with anticysticercal medications. The epidemiological criteria account for residence in or travel to areas where the disease is endemic, or having close contact with individuals infected with the tapeworm *Taenia solium*.^5^

### Study selection

Included in the study were case reports, case series, and observational studies that provided specific details on individual patients.

### Exclusion criteria

We excluded editorials, commentaries on previously published case reports, and review articles. Additionally, summaries from conferences were not considered for inclusion.

### Data extraction

The review process was conducted in two distinct stages. Initially, two independent evaluators (RK and IR) reviewed the titles and abstracts of papers. Following this, they examined the full texts of selected papers based on predefined inclusion criteria, with a third reviewer (RU) being called upon to resolve any disagreements. For the quality assessment of the included studies, a different pair of reviewers (SP and RU) were responsible, with a third reviewer (HN) stepping in as the deciding reviewer in case of unresolved disagreements.

The EndNote 21 web tool was used to identify and eliminate duplicate records, overseen by RK and HN, and assistance was sought from another reviewer in case of issues. The progress and outcomes at each stage of the review were visually represented using a PRISMA flow diagram, constructed with EndNote 21. Data collection included detailed information on patient demographics, clinical presentation, neuroimaging, treatment types, and clinical and neuroimaging outcomes of each case, jointly undertaken by three reviewers (RK, IR, and SP) with the fourth reviewer (HN) available for resolving disagreements.

The EndNote 21 software (Clarivate, based in Philadelphia, Pennsylvania, USA) was utilized for handling repeated records. All disagreements or problems encountered were settled through a mutual agreement. Additionally, a PRISMA flow diagram was created to depict the number of records gathered and evaluated at each phase, using the EndNote 21 software for assistance.

### Quality assessment

Each case was appraised based on four parameters: selection, ascertainment, causality, and reporting, as outlined by Murad MH et al.^6^

Specifically, the selection of cases was guided by whether the included cases represented the entire experience of the treating clinician. The ascertainment of diagnosis was evaluated based on the latest diagnostic criteria, cerebrospinal fluid (CSF) examination, electrophysiology, and/or neuroimaging. Causality assessment focused on the efforts made to comprehensively exclude alternative causes, and the course of illness needed to have been adequately described. The completeness of case descriptions was also assessed, ensuring that other physicians could make inferences from their experiences.

Following the criteria used by Della Gatta et al., a case report was considered “good quality” if all of the quality domains were satisfied. If three of the domains were met, the report was considered of “fair quality,” and if only two or one of the domains were satisfied, the report was deemed of “poor quality.^7^

### Data analysis

The required information was extracted and compiled into a Word table. The focused details included the first author, country of occurrence, age/sex of the patients, duration of illness, clinical manifestations, neuroimaging findings, spinal cord biopsy results, treatment administered, and the outcome of each case. Data synthesis was conducted descriptively and qualitatively. Categorical variables were depicted using frequencies and percentages, while continuous variables were reported using means or medians, along with standard deviations or ranges, as appropriate. In the final report, this data was presented in a series of tables for clear and concise representation.

## Results

Our review resulted in 163 records describing 197 patients. Figure 1 shows the PRISMA flowchart for our systematic review. Out of 197 cases evaluated, the cases were classified as having good or fair quality. (Supplementary item- 1) The PRISMA checklist has been provided as supplementary item 2. Patient related data that extracted from reports of 197 cases have been compiled in supplementary item- 3.

**Figure 1.**
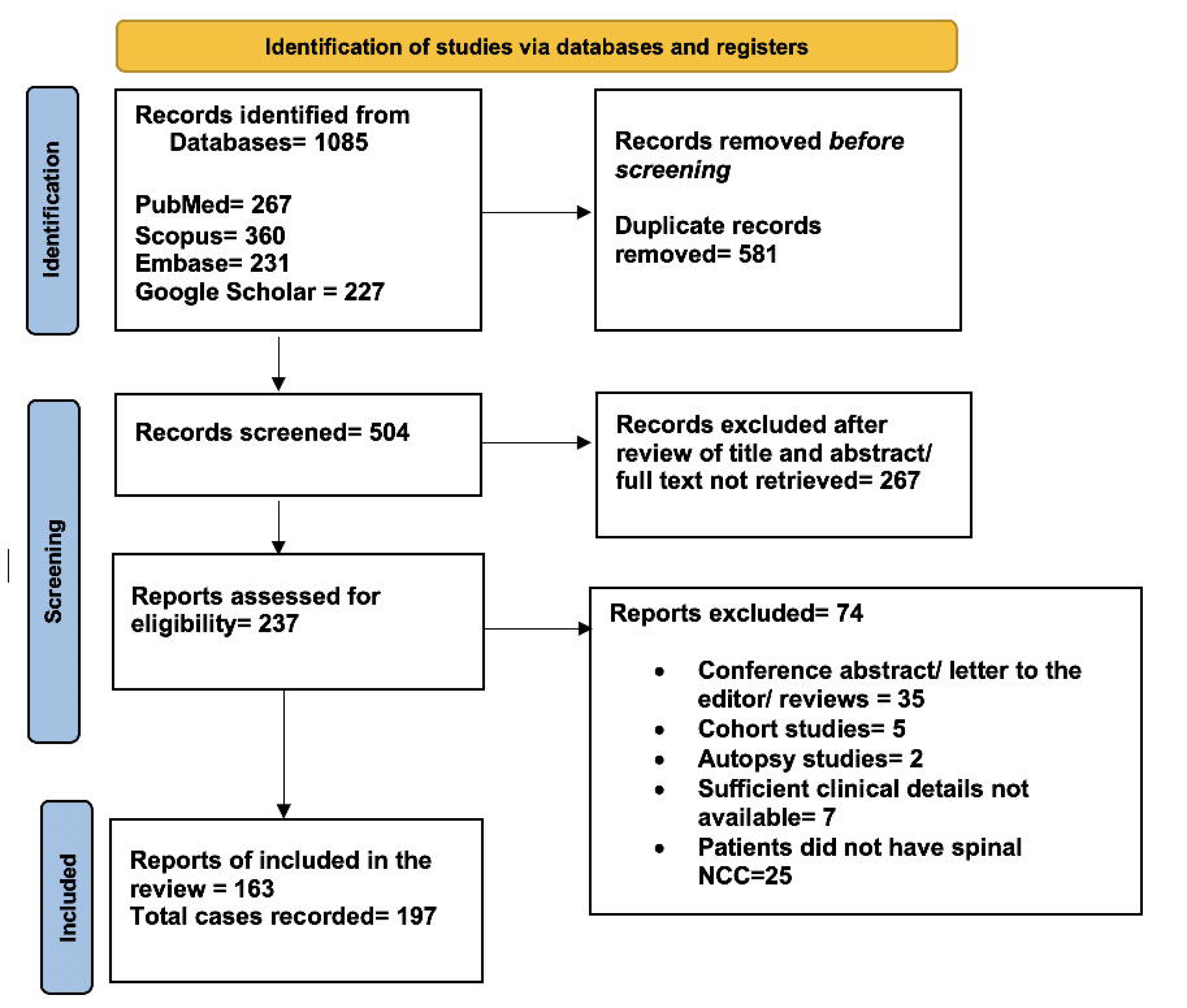
PRISMA flow diagram of the study depicts the procedure of selecting articles for the systematic review.

The mean age of the spinal neurocysticercosis patients was 35.7 years (median 40; range 5-80 years). There were a total of 74 (37.6%) females. Males were 123 (62.4%). (Table-1) The majority (60%) of cases were recorded from Asia, and the highest number of cases (64/197; 32.4%) were reported from India. (Table-1) Half of the total cases (99/197; 50.3%) presented symptoms for one week to six months. Four cases (2%) had a sudden onset. Twelve cases had symptoms for over three years. Our review noted that the majority (61%) of patients with spinal neurocysticercosis presented with paraparesis or quadriparesis. Neck pain, limb pain, and back pain were also common, affecting 41.6% of cases. In 23% of the cases (45/197), cerebrospinal fluid (CSF) abnormalities were observed, characterized by an increase in cell count and protein levels. In a few cases, low glucose and the presence of eosinophils were also noted. Among the cases, thoracic spinal cord segment involvement was most common at 40.1%, followed by cervical at 27.4%, and lumbosacral at 24.9%. There were instances of multiple or extensive involvement in 7.1% of cases. Our review revealed that intramedullary cases were the most frequent, accounting for 54.8% of the total. Intradural extramedullary cases comprised 39%, and a minimal number, 1%, were extradural. Both bone vertebral anomalies and perineural cysts were observed in 1% and 0.5% of cases, respectively. (Table-1) Our review indicates that 19.3% of the total cases had evidence of neurocysticercosis elsewhere in the body, with brain lesions being the most common at 13.7%. Intraventricular cysts were found in 1.5% of the cases and disseminated neurocysticercosis in 2.5%. Subcutaneous cysts were rare, observed in 0.5% of cases, while muscle cysts were identified in 1% of the cases. (Table-1)

The majority of spinal neurocysticercosis cases, 76.6%, presented with a cystic lesion with or without scolex. Lesions that were ring-enhancing constituted 6.6% of cases, while 15.2% had a large cyst, multiple cysts, or racemose cysts. Additionally, 14 cases were associated with surrounding arachnoiditis.

In 27% of the cases, differential diagnosis was considered. Of these, spinal cord tumors were most frequently suspected, accounting for 12.2%. Tuberculoma was considered in 3.5% of cases, while arachnoid or hydatid cysts were thought possible in 7.1%. Cavernous malformations were on the differential diagnosis list in 2% of cases. Rare considerations included multiple sclerosis and hydrocephalus at 0.5% each, and degenerative disease of the spine was considered in 1% of cases.

In the data, surgery alone was the treatment for approximately 32% of the cases. Cysticidal drugs, used with or without corticosteroids, were administered in 17.3% of cases. Corticosteroids alone were prescribed in 4.6% of cases. The combination of surgery followed by cysticidal drugs was the most preferred approach, employed in 45.2% of the cases. Within the data set, 91.9% of the cases showed improvement, while 5 cases either did not improve or resulted in death. Post-operative complications caused three deaths. (Table-1)

## Discussion

Our review indicated that the most common presenting symptom in spinal neurocysticercosis patients is paraparesis/quadriparesis and back or neck pain. The majority of patients were treated with a combination of surgical intervention and cysticidal drugs or spinal surgery alone. A smaller proportion was treated with cysticidal drugs alone. Regardless of the treatment method, most patients experienced clinical improvement. There were a few instances of post-operative complications, which led to deaths.

Spinal involvement is rare in neurocysticercosis. Any segment of the spinal cord may be involved, thoracic region is most frequently affected. our review noted that intramedullary involvement is more frequent than extramedullary involvement.

Patients generally present with quadriplegia or paraplegia depending on the location of the lesion in the spinal cord. Asymmetric neurological involvement and back pain are common in extramedullary neurocysticercosis, intramedullary lesions often present with transverse cord syndrome like the picture.^8^ If a lesion is in the lumbosacral region, patients often present with cauda-equina syndrome characterized by asymmetric lower motor neuron paraparesis, bladder disturbances, and/ or perianal saddle-shaped sensory loss.^9,10^ Spinal cord tumors, tuberculoma, and arachnoid cysts are common pre-operative diagnoses. A large racemose cyst in the subarachnoid space may cause extensive arachnoiditis.^11^ In our review, we noted that 2.5% of patients with spinal neurocysticercosis had disseminated neurocysticercosis. Ganaraja and colleagues noted disseminated neurocysticercosis in 12.5% of cases with Intramedullary spinal lesion was observed. ^12^ It is thought that the parasite can spread to the spinal cord either through the bloodstream or by directly descending along the subarachnoid space, utilizing the spinal fluid as a means of transport, in a manner influenced by gravity. Albendazole and praziquantel, often along with corticosteroids, are current medical treatments for all forms of neurocysticercosis.^13^

Clinical Practice Guidelines by the Infectious Diseases Society of America (IDSA) and the American Society of Tropical Medicine and Hygiene (ASTMH) recommended corticosteroids with or without cysticidal drugs. Intramedullary SN has traditionally been treated with laminectomy and subsequent myelotomy. There have been increasing numbers of reports of treatment with cysticidal drugs with complete recovery and disappearance of lesions on neuroimaging. For those patients with subarachnoid disease of the spine and concurrent basilar subarachnoid neurocysticercosis, medical therapy should be undertaken. Corticosteroids are critical in patients with symptomatic extramedullary spinal disease. surgical removal in the setting of severe arachnoiditis, where cysts have adhered to the sacral roots and adjacent dura can be very difficult and in some cases, complete removal of the cysts is impossible.^14^

In an earlier systematic review, data from 103 patients with spinal neurocysticercosis were reviewed. about 46% had the disease only in their spine. in the majority, the cyst was localized to an intradural extramedullary location. Almost half of the patients were treated with both surgery and cysticidal drugs, and this combination was most effective, showing better results than just surgery or medication alone. According to this systematic review, the rate of functional improvement following combined surgery and cysticidal drugs was considerably higher than that following surgery alone or medical treatment alone.^3^

Our review suggests that the use of cysticidal drugs in conjunction with corticosteroids could eliminate the need for surgery in many spinal cord neurocysticercosis patients. A randomized controlled trial is required to definitively address this question. Given the rarity of spinal cord neurocysticercosis, conducting such a study may not be feasible. Nonetheless, a thorough course of cysticidal drugs should be considered before planning surgery.

Our review has certain limitations. The variability in case presentations, imaging modalities, and responses to treatment adds a level of heterogeneity that challenges the standardization of findings. The lack of a meta-analysis further limits the study, as the descriptive approach does not allow for a quantitative data synthesis. Lastly, the potential for incomplete reporting within the sourced literature could impact the assessment of study quality and integrity.

In conclusion, surgery followed by cysticidal drugs was the most preferred treatment for spinal neurocysticercosis. Medical treatment alone was given to many patients. Clinical improvement was observed in most cases, regardless of the treatment option used. The use of cysticidal drugs could eliminate the need for surgery in many spinal neurocysticercosis patients.

## The ethical statement

No human or animal subjects were involved so ethical clearance was not taken.

## Data Availability declaration

Data is provided within the manuscript and supplementary information files.

## Supporting information

Supplementary item-1

Supplementary item-2

Supplementary item-3

---

**Table 1:** Summary of clinical features, neuroimaging characteristics and treatment outcomes in patients with spinal neurocysticercosis (n=197)

|  |  |
| --- | --- |
| **Age (in years)** | Mean= 39.2<br>Median=40<br>Mode= 40<br>Range= 5-80<br>Interquartile Range= 23 |
| **Sex** | Female= 74 (37.6%)<br>Male= 123 (62.4%) |
| **Geographical areas of reported cases (total 163 reports)** | Asia=98 (India=64 and Korea=17) (60.1%)<br>Europe=10 (7.3%)<br>Africa=2 (1.2%)<br>North America=29 (USA=25) (17.8%)<br>South America=22 (Brazil=18) (13.%%)<br>Australia=2 (1.2%) |
| **Duration of illness** | NA=26 (13.1%)<br>Sudden= 4 (2%)<br>Up to 1 week=15 (7.6%)<br>>7 days to one month= 40 (20.3%)<br>>One month to 6 months=59 (30%)<br>> 6 months to 1 year=21 (10.7%)<br>> 1 year to 3 years=24 (12.2%)<br>> 3 years to 10 years= 6 (3%)<br>> 10 years=2 (1%) |
| **Clinical features** | Neck pain/limb pain/back pain= 82 (41.6%)<br>Headache= 17 (8.6%)<br>Paraparesis/ quadriparesis= 120 (61%)<br>Cauda-equina syndrome= 8 (4%)<br>Difficulty in walking= 6(3%)<br>Unilateral limb weakness= 10 (5.1%)<br>Seizures=7 (3.6%) |
| **CSF** | Abnormal= 45 (22.8%)<br>NA/normal=152 (77.2%) |
| **Spinal segment involvement** | Cervical= 54 (27.4%)<br>Thoracic = 79 (40.1%)<br>Lumbo-sacral= 49 (24.9%)<br>Multiple/ extensive=14 (7.1%)<br>Bony vertebral=1 (0.5%) |
| **Type of spinal cord involvement** | NA=6 (3%)<br>Intradural extramedullary=77 (39%)<br>Intramedullary=108 (54.8%)<br>Extradural=2 (1%)<br>Bone vertebral=2 (1%)<br>Perineural cysts=1 (0.5%)<br>Conus terminallis=1 (0.5%) |
| **NCC lesions elsewhere** | Total= 38 (19.3%)<br>Brain lesions= 27 (13.7%) |
|  | Intraventricular cysts= 3 (1.5%)<br>Disseminated neurocysticercosis=5 (2.5%)<br>Subcutaneous cysts=1(0.5%)<br>Muscle cysts= 2(1%) |
| **Lesion characteristics** | A ring- enhancing lesion= 13 (6.6%)<br>A large cyst, multiple cysts or racemose cysts=30 (15.2%)<br>Cystic lesion with or without scolex=151 (76.6%)<br>Only arachnoiditis= 1(0.5%)<br>NA=1(0.5%)<br>Not- well defined=1(0.5%)<br>Surrounding arachnoiditis= 14 |
| **Differential diagnosis** | The cases where a differential diagnosis was considered= 53 (27%)<br>Common differential diagnosis were<br>Tuberculoma= 7 (3.5%)<br>Spinal cord tumor= 24 (12.2%)<br>Arachnoid cysts/hydatid cysts= 14 (7.1%)<br>Cavernous malformation= 4(2%)<br>Multiple sclerosis=1(0.5%)<br>Degenerative disease of spine=2(1%)<br>Hydrocephalus-1(0.5%) |
| **Treatment modalities** | Surgery only= 63 (32%)<br>Cysticidal drugs with or without Corticosteroids = 34 (17.3%)<br>Corticosteroids only =9 (4.6%)<br>Surgery followed by cysticidal drugs= 89 (45.2%) |
| **Outcome** | NA= 11 (5.6%)<br>Died/ not improved=5 (2.5%)<br>Improved= 181 (91.9%) |

## References

1. World Health Organization. Taeniasis/cysticercosis. Key facts 11 January 2022. Downloaded from https://www.who.int/news-room/fact-sheets/detail/taeniasis-cysticercosis. Assessed on 21 January 2024.

2. World Health Organization. WHO guidelines on the management of Taenia solium neurocysticercosis. Technical document. 2 September 2021. Downloaded from https://www.who.int/publications/i/item/9789240032231. Assessed on 21 January, 2024.

3. Barrie U, Badejo O, Aoun SG, Adeyemo E, Moler N, Christian ZK, et al. Systematic Review and Meta-Analysis of Management Strategies and Outcomes in Adult Spinal Neurocysticercosis. World Neurosurg. 2020;138:504–511.e8.

4. Garg RK, Suresh V, Rizvi I, Nigam H, Pandey S. Treatment outcome in patients with spinal neurocysticercosis: a systematic review of published cases and case series. PROSPERO 2024 CRD42024496957 Available from: https://www.crd.york.ac.uk/prospero/display_record.php?ID=CRD42024496957.

5. Del Brutto OH, Nash TE, White AC Jr, Rajshekhar V, Wilkins PP, Singh G, et al. Revised diagnostic criteria for neurocysticercosis. J Neurol Sci. 2017;372:202–210.

6. Murad MH, Sultan S, Haffar S, Bazerbachi F. Methodological quality and synthesis of case series and case reports. BMJ Evid Based Med. 2018;23:60–63.

7. Della Gatta AN, Rizzo R, Pilu G, Simonazzi G. Coronavirus disease 2019 during pregnancy: a systematic review of reported cases. Am J Obstet Gynecol. 2020;223:36–41.

8. Del Brutto OH, Garcia HH. Intramedullary cysticercosis of the spinal cord: a review of patients evaluated with MRI. J Neurol Sci. 2013;331:114–7.

9. Dhar A, Dua S, Singh H. Isolated Intramedullary Lumbar Spine Neurocysticercosis: A Rare Occurrence and Review of Literature. Surg J (N Y). 2021;7:e327–e336.

10. Zhang S, Hu Y, Li Z, Zhao L, Wang Z. Lumbar spinal intradural neurocysticercosis: A case report. Exp Ther Med. 2017;13(6):3591–3593.

11. Yacoub HA, Goldstein I, El-Ghanem M, Sharer L, Souayah N. Spinal racemose cysticercosis: case report and review. Hosp Pract (1995). 2017;45(3):99-103.

12. Ganaraja HV, Mahadevan A, Saini J, Nalini A, Pal PK, Satishchandra P, et al. Disseminated Cysticercosis in Indian Scenario - Experience from a Teaching University Hospital. Neurol India. 2022;70(3):1032-1040.

13. Gripper LB, Welburn SC. Neurocysticercosis infection and disease-A review. Acta Trop. 2017;166:218–224.

14. White AC Jr, Coyle CM, Rajshekhar V, Singh G, Hauser WA, Mohanty A, et al. Diagnosis and Treatment of Neurocysticercosis: 2017 Clinical Practice Guidelines by the Infectious Diseases Society of America (IDSA) and the American Society of Tropical Medicine and Hygiene (ASTMH). Am J Trop Med Hyg. 2018;98(4):945-966.

