## Supplementary item-2 for "Treatment Outcome in Patients With Spinal Neurocysticercosis: A Systematic Review of Published Cases and Case Series"

**Supplementary table-2: Evaluation of the methodological quality of case reports and case series**

| **Reference** | **Does the patient represent the whole experience of the investigator** | **Was the exposure adequately ascertained?** | **Was the outcome adequately ascertained?** | **Were other alternative causes that may explain the observation ruled out?** | **Was there a challenge and/or re-challenge phenomenon?** | **Was there a dose-response effect?** | **Was follow-up long enough for outcomes to occur?** | **Is the case(s) described with sufficient details to allow practitioners make inferences related to their own practice?** | **Score** |
| --- | --- | --- | --- | --- | --- | --- | --- | --- | --- |
| **Vijayan et al 2023** | **Yes** | **Yes** | **Yes** | **Yes** | **NA** | **Yes** | **Yes** | **Yes** | **7** |
| **Tao et al 2023** | **Yes** | **Yes** | **Yes** | **Yes** | **NA** | **Yes** | **Yes** | **Yes** | **7** |
| **Pedrosa et al 2023** | **Yes** | **Yes** | **Yes** | **Yes** | **NA** | **Yes** | **Yes** | **Yes** | **7** |
| **Manh et al 2023** | **Yes** | **Yes** | **Yes** | **Yes** | **NA** | **Yes** | **Yes** | **Yes** | **7** |
| **Machado et al 2023** | **Yes** | **Yes** | **Yes** | **Yes** | **NA** | **Yes** | **Yes** | **Yes** | **7** |
| **Lama et al 2023** | **Yes** | **Yes** | **Yes** | **Yes** | **NA** | **Yes** | **Yes** | **Yes** | **7** |
| **Canales et al 2023** | **Yes** | **Yes** | **Yes** | **Yes** | **NA** | **Yes** | **Yes** | **Yes** | **7** |
| **Almeida et al 2023** | **Yes** | **Yes** | **Yes** | **Yes** | **NA** | **Yes** | **Yes** | **Yes** | **7** |
| **Zheng et al 2022** | **Yes** | **Yes** | **Yes** | **Yes** | **NA** | **Yes** | **Yes** | **Yes** | **7** |
| **Yang et al 2022** | **Yes** | **Yes** | **Yes** | **Yes** | **NA** | **Yes** | **Yes** | **Yes** | **7** |
|  | **Yes** | **Yes** | **Yes** | **Yes** | **NA** | **Yes** | **Yes** | **Yes** | **7** |
|  | **Yes** | **Yes** | **Yes** | **Yes** | **NA** | **Yes** | **Yes** | **Yes** | **7** |
|  | **Yes** | **Yes** | **Yes** | **Yes** | **NA** | **Yes** | **Yes** | **Yes** | **7** |
|  | **Yes** | **Yes** | **Yes** | **Yes** | **NA** | **Yes** | **Yes** | **Yes** | **7** |
|  | **Yes** | **Yes** | **Yes** | **Yes** | **NA** | **Yes** | **Yes** | **Yes** | **7** |
|  | **Yes** | **Yes** | **Yes** | **Yes** | **NA** | **Yes** | **Yes** | **Yes** | **7** |
| **Solanki et al 2022** | **Yes** | **Yes** | **Yes** | **Yes** | **NA** | **Yes** | **Yes** | **Yes** | **7** |
| **Sihag et al 2022** | **Yes** | **Yes** | **Yes** | **Yes** | **NA** | **Yes** | **Yes** | **Yes** | **7** |
| **Roy et al 2022** | **Yes** | **Yes** | **Yes** | **Yes** | **NA** | **Yes** | **Yes** | **Yes** | **7** |
| **Kus et al 2022** | **Yes** | **Yes** | **Yes** | **Yes** | **NA** | **Yes** | **Yes** | **Yes** | **7** |
| **Kumar et al 2022** | **Yes** | **Yes** | **Yes** | **Yes** | **NA** | **Yes** | **Yes** | **Yes** | **7** |
| **Kim et al 2022** | **Yes** | **Yes** | **Yes** | **Yes** | **NA** | **Yes** | **Yes** | **Yes** | **7** |
| **Gorjian and Ricks 2022** | **Yes** | **Yes** | **Yes** | **Yes** | **NA** | **Yes** | **Yes** | **Yes** | **7** |
| **Andino et al 2022** | **Yes** | **Yes** | **Yes** | **Yes** | **NA** | **Yes** | **Yes** | **Yes** | **7** |
| **Chenyu et al 2022** | **Yes** | **Yes** | **Yes** | **Yes** | **NA** | **Yes** | **Yes** | **Yes** | **7** |
| **Vadher et al 2021** | **Yes** | **Yes** | **Yes** | **Yes** | **NA** | **Yes** | **Yes** | **Yes** | **7** |
| **Rajbhandari et al 2021** | **Yes** | **Yes** | **Yes** | **Yes** | **NA** | **Yes** | **Yes** | **Yes** | **7** |
| **Radhakrishnan 2021** | **Yes** | **Yes** | **Yes** | **Yes** | **NA** | **Yes** | **Yes** | **Yes** | **7** |
| **Mediratta 2021** | **Yes** | **Yes** | **Yes** | **Yes** | **NA** | **Yes** | **Yes** | **Yes** | **7** |
| **Lahiri et al 2021** | **Yes** | **Yes** | **Yes** | **Yes** | **NA** | **Yes** | **Yes** | **Yes** | **7** |
| **Garg et al 2021** | **Yes** | **Yes** | **Yes** | **Yes** | **NA** | **Yes** | **Yes** | **Yes** | **7** |
| **Dhar et al 2021** | **Yes** | **Yes** | **Yes** | **Yes** | **NA** | **Yes** | **Yes** | **Yes** | **7** |
| **Chandrakanth et al 2021** | **Yes** | **Yes** | **Yes** | **Yes** | **NA** | **Yes** | **Yes** | **Yes** | **7** |
| **Yu et al 2020** | **Yes** | **Yes** | **Yes** | **Yes** | **NA** | **Yes** | **Yes** | **Yes** | **7** |
| **Walia et al 2020** | **Yes** | **Yes** | **Yes** | **Yes** | **NA** | **Yes** | **Yes** | **Yes** | **7** |
| **Jobanputra et al 2020** | **Yes** | **Yes** | **Yes** | **Yes** | **NA** | **Yes** | **Yes** | **Yes** | **7** |
| **Gawande et al 2020** | **Yes** | **Yes** | **Yes** | **Yes** | **NA** | **Yes** | **NO** | **NO** | **5** |
| **Asín et al 2020** | **Yes** | **Yes** | **Yes** | **Yes** | **NA** | **Yes** | **Yes** | **Yes** | **7** |
| **Ansari et al 2020** | **Yes** | **Yes** | **Yes** | **Yes** | **NA** | **Yes** | **Yes** | **Yes** | **7** |
| **Torres-Corzo et al 2019** | **Yes** | **Yes** | **Yes** | **Yes** | **NA** | **Yes** | **Yes** | **Yes** | **7** |
|  | **Yes** | **Yes** | **Yes** | **Yes** | **NA** | **Yes** | **Yes** | **Yes** | **7** |
|  | **Yes** | **Yes** | **Yes** | **Yes** | **NA** | **Yes** | **Yes** | **Yes** | **7** |
| **Lopez et al 2019** | **Yes** | **Yes** | **Yes** | **Yes** | **NA** | **Yes** | **Yes** | **Yes** | **7** |
| **Shashidhar et al 2018** | **Yes** | **Yes** | **Yes** | **Yes** | **NA** | **Yes** | **Yes** | **Yes** | **7** |
| **Phuyal et al 2018** | **Yes** | **Yes** | **Yes** | **Yes** | **NA** | **Yes** | **Yes** | **Yes** | **7** |
| **Maste et al 2018** | **Yes** | **Yes** | **Yes** | **Yes** | **NA** | **Yes** | **Yes** | **Yes** | **7** |
| **Jeong et al 2018** | **Yes** | **Yes** | **Yes** | **Yes** | **NA** | **Yes** | **Yes** | **Yes** | **7** |
| **Agarwal et al 2018** | **Yes** | **Yes** | **Yes** | **Yes** | **NA** | **Yes** | **Yes** | **Yes** | **7** |
| **Giri et al 2018** | **Yes** | **Yes** | **Yes** | **Yes** | **NA** | **Yes** | **Yes** | **Yes** | **7** |
| **Almeida et al 2018** | **Yes** | **Yes** | **Yes** | **Yes** | **NA** | **Yes** | **Yes** | **NO** | **6** |
| **Zhang et al 2017** | **Yes** | **Yes** | **Yes** | **Yes** | **NA** | **Yes** | **Yes** | **Yes** | **7** |
| **Yadav et al 2017** | **Yes** | **Yes** | **Yes** | **Yes** | **NA** | **Yes** | **Yes** | **Yes** | **7** |
| **Yacoub et al 2017** | **Yes** | **Yes** | **Yes** | **Yes** | **NA** | **Yes** | **Yes** | **Yes** | **7** |
| **Vetrivel et al 2017** | **Yes** | **Yes** | **Yes** | **Yes** | **NA** | **Yes** | **Yes** | **Yes** | **7** |
| **Sharma et al 2017** | **Yes** | **Yes** | **Yes** | **Yes** | **NA** | **Yes** | **Yes** | **Yes** | **7** |
| **Santos et al 2017** | **Yes** | **Yes** | **Yes** | **Yes** | **NA** | **Yes** | **Yes** | **Yes** | **7** |
| **Ranjan et al 2017** | **Yes** | **Yes** | **Yes** | **Yes** | **NA** | **Yes** | **Yes** | **Yes** | **7** |
| **Pal et al 2017** | **Yes** | **Yes** | **Yes** | **Yes** | **NA** | **Yes** | **Yes** | **Yes** | **7** |
| **Mesquita Filho et al 2017** | **Yes** | **Yes** | **Yes** | **Yes** | **NA** | **Yes** | **Yes** | **Yes** | **7** |
| **Hedaoo et al 2017** | **Yes** | **Yes** | **Yes** | **Yes** | **NA** | **Yes** | **Yes** | **Yes** | **7** |
| **Hansberry et al 2017** | **Yes** | **Yes** | **Yes** | **Yes** | **NA** | **Yes** | **Yes** | **Yes** | **7** |
| **Datta et al 2017** | **Yes** | **Yes** | **Yes** | **Yes** | **NA** | **Yes** | **Yes** | **Yes** | **7** |
|  | **Yes** | **Yes** | **Yes** | **Yes** | **NA** | **Yes** | **Yes** | **Yes** | **7** |
|  | **Yes** | **Yes** | **Yes** | **Yes** | **NA** | **NO** | **NO** | **NO** | **4** |
| **Bansal et al 2017** | **Yes** | **Yes** | **Yes** | **Yes** | **NA** | **Yes** | **Yes** | **Yes** | **7** |
| **Torous and Darras 2016** | **Yes** | **Yes** | **Yes** | **Yes** | **NA** | **Yes** | **Yes** | **Yes** | **7** |
| **Pant et al 2016** | **Yes** | **Yes** | **Yes** | **Yes** | **NA** | **Yes** | **Yes** | **Yes** | **7** |
|  | **Yes** | **Yes** | **Yes** | **Yes** | **NA** | **Yes** | **Yes** | **Yes** | **7** |
| **Pratap Kumar and Ravi 2016** | **Yes** | **Yes** | **Yes** | **Yes** | **NA** | **Yes** | **Yes** | **Yes** | **7** |
| **Mewada and Srivastava 2016** | **Yes** | **Yes** | **Yes** | **Yes** | **NA** | **Yes** | **Yes** | **Yes** | **7** |
| **Wang and Huang 2015** | **Yes** | **Yes** | **Yes** | **Yes** | **NA** | **Yes** | **Yes** | **Yes** | **7** |
| **Veiga et al 2015** | **Yes** | **Yes** | **Yes** | **Yes** | **NA** | **Yes** | **Yes** | **Yes** | **7** |
| **Valsangkar et al 2015** | **Yes** | **Yes** | **Yes** | **Yes** | **NA** | **Yes** | **Yes** | **Yes** | **7** |
| **Salazar Noguera et al 2015** | **Yes** | **Yes** | **Yes** | **Yes** | **NA** | **Yes** | **Yes** | **Yes** | **7** |
| **Ruschel et al 2015** | **Yes** | **Yes** | **Yes** | **Yes** | **NA** | **Yes** | **Yes** | **Yes** | **7** |
| **Hackius et al 2015** | **Yes** | **Yes** | **Yes** | **Yes** | **NA** | **Yes** | **Yes** | **Yes** | **7** |
| **Ganesan et al 2015** | **Yes** | **Yes** | **Yes** | **Yes** | **NA** | **Yes** | **Yes** | **Yes** | **7** |
| **Chaurasia et al 2015** | **Yes** | **Yes** | **Yes** | **Yes** | **NA** | **Yes** | **Yes** | **Yes** | **7** |
| **Bhardwaj 2015** | **Yes** | **Yes** | **Yes** | **Yes** | **NA** | **Yes** | **NO** | **NO** | **5** |
| **Yoo et al 2014** | **Yes** | **Yes** | **Yes** | **Yes** | **NA** | **Yes** | **Yes** | **Yes** | **7** |
| **Verma et al 2014** | **Yes** | **Yes** | **Yes** | **Yes** | **NA** | **Yes** | **Yes** | **Yes** | **7** |
| **Qazi et al 2014** | **Yes** | **Yes** | **Yes** | **Yes** | **NA** | **Yes** | **Yes** | **Yes** | **7** |
| **Kim et al 2014** | **Yes** | **Yes** | **Yes** | **Yes** | **NA** | **Yes** | **Yes** | **Yes** | **7** |
| **Kim et al 2014** | **Yes** | **Yes** | **Yes** | **Yes** | **NA** | **Yes** | **Yes** | **Yes** | **7** |
| **Han et al 2014** | **Yes** | **Yes** | **Yes** | **Yes** | **NA** | **Yes** | **Yes** | **Yes** | **7** |
| **Ahmed and Paul 2014** | **Yes** | **Yes** | **Yes** | **Yes** | **NA** | **Yes** | **Yes** | **Yes** | **7** |
| **Abarrategui Yagüe et al 2014** | **Yes** | **Yes** | **Yes** | **Yes** | **NA** | **Yes** | **Yes** | **Yes** | **7** |
| **Iacoangeli et al 2013** | **Yes** | **Yes** | **Yes** | **Yes** | **NA** | **Yes** | **Yes** | **Yes** | **7** |
| **Furtado et al 2013** | **Yes** | **Yes** | **Yes** | **Yes** | **NA** | **Yes** | **Yes** | **Yes** | **7** |
| **De Feo et al 2013** | **Yes** | **Yes** | **Yes** | **Yes** | **NA** | **Yes** | **Yes** | **Yes** | **7** |
| **Chandramohan et al 2013** | **Yes** | **Yes** | **Yes** | **Yes** | **NA** | **Yes** | **Yes** | **Yes** | **7** |
| **Araujo et al 2013** | **Yes** | **Yes** | **Yes** | **Yes** | **NA** | **Yes** | **Yes** | **Yes** | **7** |
| **Shin et al 2012** | **Yes** | **Yes** | **Yes** | **Yes** | **NA** | **Yes** | **Yes** | **Yes** | **7** |
| **Rice and Perera 2012** | **Yes** | **Yes** | **Yes** | **Yes** | **NA** | **Yes** | **Yes** | **Yes** | **7** |
| **Naguib et al 2012** | **Yes** | **Yes** | **Yes** | **Yes** | **NA** | **NO** | **NO** | **NO** | **4** |
| **Motsepe and Ackerman 2012** | **Yes** | **Yes** | **Yes** | **Yes** | **NA** | **Yes** | **Yes** | **Yes** | **7** |
| **Kapu et al 2012** | **Yes** | **Yes** | **Yes** | **Yes** | **NA** | **Yes** | **Yes** | **Yes** | **7** |
| **Jain et al 2012** | **Yes** | **Yes** | **Yes** | **Yes** | **NA** | **Yes** | **Yes** | **Yes** | **7** |
| **Bhowmik et al 2012** | **Yes** | **Yes** | **Yes** | **Yes** | **NA** | **Yes** | **Yes** | **Yes** | **7** |
| **Agale et al 2012** | **Yes** | **Yes** | **Yes** | **Yes** | **NA** | **Yes** | **Yes** | **Yes** | **7** |
| **Vij et al 2011** | **Yes** | **Yes** | **Yes** | **Yes** | **NA** | **Yes** | **Yes** | **Yes** | **7** |
| **Seo et al 2011** | **Yes** | **Yes** | **Yes** | **Yes** | **NA** | **Yes** | **Yes** | **Yes** | **7** |
| **Qi et al 2011** | **Yes** | **Yes** | **Yes** | **Yes** | **NA** | **Yes** | **Yes** | **Yes** | **7** |
| **Park et al 2011** | **Yes** | **Yes** | **Yes** | **Yes** | **NA** | **Yes** | **Yes** | **Yes** | **7** |
| **Lambertucci et al 2011** | **Yes** | **Yes** | **Yes** | **Yes** | **NA** | **Yes** | **Yes** | **Yes** | **7** |
| **Jongwutiwes et al 2011** | **Yes** | **Yes** | **Yes** | **Yes** | **NA** | **Yes** | **Yes** | **Yes** | **7** |
| **Heredia Mo et al 2011** | **Yes** | **Yes** | **Yes** | **Yes** | **NA** | **Yes** | **Yes** | **Yes** | **7** |
| **Azfar et al 2011** | **Yes** | **Yes** | **Yes** | **Yes** | **NA** | **Yes** | **Yes** | **Yes** | **7** |
| **Ahuja et al 2011** | **Yes** | **Yes** | **Yes** | **Yes** | **NA** | **Yes** | **Yes** | **Yes** | **7** |
| **Lin et al 2010** | **Yes** | **Yes** | **Yes** | **Yes** | **NA** | **Yes** | **Yes** | **Yes** | **7** |
| **Lim et al 2010** | **Yes** | **Yes** | **Yes** | **Yes** | **NA** | **Yes** | **Yes** | **Yes** | **7** |
| **Kumar et al 2010** | **Yes** | **Yes** | **Yes** | **Yes** | **NA** | **Yes** | **Yes** | **Yes** | **7** |
| **Jang et al 2010** | **Yes** | **Yes** | **Yes** | **Yes** | **NA** | **Yes** | **Yes** | **Yes** | **7** |
| **Gonçalves et al 2010** | **Yes** | **Yes** | **Yes** | **Yes** | **NA** | **Yes** | **Yes** | **Yes** | **7** |
| **Dhillon et al 2010** | **Yes** | **Yes** | **Yes** | **Yes** | **NA** | **Yes** | **Yes** | **Yes** | **7** |
| **Choi et al 2010** | **Yes** | **Yes** | **Yes** | **Yes** | **NA** | **Yes** | **Yes** | **Yes** | **7** |
| **Shin and Shin 2009** | **Yes** | **Yes** | **Yes** | **Yes** | **NA** | **Yes** | **Yes** | **Yes** | **7** |
| **Gupta et al 2009** | **Yes** | **Yes** | **Yes** | **Yes** | **NA** | **Yes** | **Yes** | **Yes** | **7** |
| **Chhiber et al 2009** | **Yes** | **Yes** | **Yes** | **Yes** | **NA** | **Yes** | **Yes** | **Yes** | **7** |
| **Mohapatra et al 2008** | **Yes** | **Yes** | **Yes** | **Yes** | **NA** | **Yes** | **Yes** | **Yes** | **7** |
| **Kasliwal et al 2008** | **Yes** | **Yes** | **Yes** | **Yes** | **NA** | **Yes** | **Yes** | **Yes** | **7** |
| **Izci et al 2008** | **Yes** | **Yes** | **Yes** | **Yes** | **NA** | **Yes** | **Yes** | **Yes** | **7** |
| **Edwards et al 2008** | **Yes** | **Yes** | **Yes** | **Yes** | **NA** | **Yes** | **Yes** | **Yes** | **7** |
| **Agrawal et al 2008** | **Yes** | **Yes** | **Yes** | **Yes** | **NA** | **Yes** | **Yes** | **Yes** | **7** |
| **Paterakis et al 2007** | **Yes** | **Yes** | **Yes** | **Yes** | **NA** | **Yes** | **Yes** | **Yes** | **7** |
| **Ahmad and Sharma 2007** | **Yes** | **Yes** | **Yes** | **Yes** | **NA** | **Yes** | **Yes** | **Yes** | **7** |
|  | **Yes** | **Yes** | **Yes** | **Yes** | **NA** | **Yes** | **Yes** | **Yes** | **7** |
| **Rossi et al 2006** | **Yes** | **Yes** | **Yes** | **Yes** | **NA** | **Yes** | **NO** | **NO** | **5** |
| **Guedes-Corrêa et al 2006** | **Yes** | **Yes** | **Yes** | **Yes** | **NA** | **Yes** | **Yes** | **Yes** | **7** |
| **Kim and Lee 2005** | **Yes** | **Yes** | **Yes** | **Yes** | **NA** | **Yes** | **Yes** | **Yes** | **7** |
| **Torabi et al 2004** | **Yes** | **Yes** | **Yes** | **Yes** | **NA** | **Yes** | **Yes** | **Yes** | **7** |
| **Singh and Sahai 2004** | **Yes** | **Yes** | **Yes** | **Yes** | **NA** | **Yes** | **Yes** | **Yes** | **7** |
| **Jarupant et al 2004** | **Yes** | **Yes** | **Yes** | **Yes** | **NA** | **Yes** | **Yes** | **Yes** | **7** |
| **Delobe et al 2004** | **Yes** | **Yes** | **Yes** | **Yes** | **NA** | **Yes** | **Yes** | **Yes** | **7** |
| **Jang et al 2003** | **Yes** | **Yes** | **Yes** | **Yes** | **NA** | **Yes** | **Yes** | **Yes** | **7** |
| **Costa Junior et al 2003** | **Yes** | **Yes** | **Yes** | **Yes** | **NA** | **Yes** | **Yes** | **Yes** | **7** |
| **Sheehan et al 2002** | **Yes** | **Yes** | **Yes** | **Yes** | **NA** | **Yes** | **Yes** | **Yes** | **7** |
| **Yoon et al 2002** | **Yes** | **Yes** | **Yes** | **Yes** | **NA** | **Yes** | **Yes** | **Yes** | **7** |
| **Muzumdar et al 2002** | **Yes** | **Yes** | **Yes** | **Yes** | **NA** | **Yes** | **Yes** | **Yes** | **7** |
|  | **Yes** | **Yes** | **Yes** | **Yes** | **NA** | **Yes** | **Yes** | **Yes** | **7** |
| **Alsina et al 2002** | **Yes** | **Yes** | **Yes** | **Yes** | **NA** | **Yes** | **Yes** | **Yes** | **7** |
|  | **Yes** | **Yes** | **Yes** | **Yes** | **NA** | **Yes** | **Yes** | **Yes** | **7** |
|  | **Yes** | **Yes** | **Yes** | **Yes** | **NA** | **Yes** | **Yes** | **Yes** | **7** |
|  | **Yes** | **Yes** | **Yes** | **Yes** | **NA** | **Yes** | **Yes** | **Yes** | **7** |
|  | **Yes** | **Yes** | **Yes** | **Yes** | **NA** | **Yes** | **Yes** | **Yes** | **7** |
|  | **Yes** | **Yes** | **Yes** | **Yes** | **NA** | **Yes** | **Yes** | **Yes** | **7** |
| **Parmar et al 2001** | **Yes** | **Yes** | **Yes** | **Yes** | **NA** | **Yes** | **Yes** | **Yes** | **7** |
|  | **Yes** | **Yes** | **Yes** | **Yes** | **NA** | **Yes** | **Yes** | **Yes** | **7** |
|  | **Yes** | **Yes** | **Yes** | **Yes** | **NA** | **Yes** | **Yes** | **Yes** | **7** |
|  | **Yes** | **Yes** | **Yes** | **Yes** | **NA** | **Yes** | **Yes** | **Yes** | **7** |
|  | **Yes** | **Yes** | **Yes** | **Yes** | **NA** | **Yes** | **Yes** | **Yes** | **7** |
|  | **Yes** | **Yes** | **Yes** | **Yes** | **NA** | **Yes** | **Yes** | **Yes** | **7** |
| **Mathuriya et al 2001** | **Yes** | **Yes** | **Yes** | **Yes** | **NA** | **Yes** | **Yes** | **Yes** | **7** |
|  | **Yes** | **Yes** | **Yes** | **Yes** | **NA** | **Yes** | **Yes** | **Yes** | **7** |
|  | **Yes** | **Yes** | **Yes** | **Yes** | **NA** | **Yes** | **Yes** | **Yes** | **7** |
| **Homans et al 2001** | **Yes** | **Yes** | **Yes** | **Yes** | **NA** | **Yes** | **Yes** | **Yes** | **7** |
| **Sahoo 2000** | **Yes** | **Yes** | **Yes** | **Yes** | **NA** | **Yes** | **Yes** | **Yes** | **7** |
| **Gaur et al 2000** | **Yes** | **Yes** | **Yes** | **Yes** | **NA** | **Yes** | **Yes** | **Yes** | **7** |
|  | **Yes** | **Yes** | **Yes** | **Yes** | **NA** | **Yes** | **Yes** | **Yes** | **7** |
| **Dantas et al 1999** | **Yes** | **Yes** | **Yes** | **Yes** | **NA** | **Yes** | **Yes** | **Yes** | **7** |
| **Ciftci et al 1999** | **Yes** | **Yes** | **Yes** | **Yes** | **NA** | **Yes** | **NO** | **NO** | **5** |
| **Rosahl and Samii 1998** | **Yes** | **Yes** | **Yes** | **Yes** | **NA** | **Yes** | **NO** | **NO** | **5** |
| **Mohanty et al 1998** | **Yes** | **Yes** | **Yes** | **Yes** | **NA** | **Yes** | **Yes** | **Yes** | **7** |
| **Lau et al 1998** | **Yes** | **Yes** | **Yes** | **Yes** | **NA** | **Yes** | **Yes** | **Yes** | **7** |
| **Garg and Nag 1998** | **Yes** | **Yes** | **Yes** | **Yes** | **NA** | **Yes** | **Yes** | **Yes** | **7** |
|  | **Yes** | **Yes** | **Yes** | **Yes** | **NA** | **Yes** | **Yes** | **Yes** | **7** |
| **Escobar et al 1998** | **Yes** | **Yes** | **Yes** | **Yes** | **NA** | **Yes** | **Yes** | **NO** | **6** |
| **Davies et al 1996** | **Yes** | **Yes** | **Yes** | **Yes** | **NA** | **Yes** | **Yes** | **Yes** | **7** |
| **Corral et al 1996** | **Yes** | **Yes** | **Yes** | **Yes** | **NA** | **Yes** | **Yes** | **Yes** | **7** |
| **Prasathapong 1995** | **Yes** | **Yes** | **Yes** | **Yes** | **NA** | **Yes** | **Yes** | **Yes** | **7** |
| **Kim et al 1995** | **Yes** | **Yes** | **Yes** | **Yes** | **NA** | **Yes** | **Yes** | **Yes** | **7** |
|  | **Yes** | **Yes** | **Yes** | **Yes** | **NA** | **Yes** | **NO** | **NO** | **5** |
|  | **Yes** | **Yes** | **Yes** | **Yes** | **NA** | **Yes** | **Yes** | **Yes** | **7** |
|  | **Yes** | **Yes** | **Yes** | **Yes** | **NA** | **Yes** | **Yes** | **Yes** | **7** |
| **Gallani et al 1992** | **Yes** | **Yes** | **Yes** | **Yes** | **NA** | **Yes** | **Yes** | **Yes** | **7** |
|  | **Yes** | **Yes** | **Yes** | **Yes** | **NA** | **Yes** | **Yes** | **Yes** | **7** |
| **Bandres et al 1992** | **Yes** | **Yes** | **Yes** | **Yes** | **NA** | **Yes** | **Yes** | **Yes** | **7** |
| **Palasis and Drevelengas 1991** | **Yes** | **Yes** | **Yes** | **Yes** | **NA** | **Yes** | **Yes** | **Yes** | **7** |
| **Venkataramana et al 1989** | **Yes** | **Yes** | **Yes** | **Yes** | **NA** | **Yes** | **Yes** | **Yes** | **7** |
|  | **Yes** | **Yes** | **Yes** | **Yes** | **NA** | **Yes** | **Yes** | **Yes** | **7** |
| **Sperlescu et al 1989** | **Yes** | **Yes** | **Yes** | **Yes** | **NA** | **Yes** | **Yes** | **Yes** | **7** |
|  | **Yes** | **Yes** | **Yes** | **Yes** | **NA** | **Yes** | **Yes** | **Yes** | **7** |
| **Vlok and Wells 1988** | **Yes** | **Yes** | **Yes** | **Yes** | **NA** | **Yes** | **Yes** | **Yes** | **7** |
| **Castillo et al 1988** | **Yes** | **Yes** | **Yes** | **Yes** | **NA** | **Yes** | **Yes** | **Yes** | **7** |
| **Sharma et al 1987** | **Yes** | **Yes** | **Yes** | **Yes** | **NA** | **Yes** | **Yes** | **Yes** | **7** |
| **Savoiardo et al 1986** | **Yes** | **Yes** | **Yes** | **Yes** | **NA** | **Yes** | **Yes** | **Yes** | **7** |
| **Holtzman et al 1986** | **Yes** | **Yes** | **Yes** | **Yes** | **NA** | **Yes** | **Yes** | **Yes** | **7** |
| **Kim et al 1985** | **Yes** | **Yes** | **Yes** | **Yes** | **NA** | **Yes** | **NO** | **NO** | **5** |
| **McDonald et al 1979** | **Yes** | **Yes** | **Yes** | **Yes** | **NA** | **Yes** | **Yes** | **Yes** | **7** |
| **Akiguchi et al 1979** | **Yes** | **Yes** | **Yes** | **Yes** | **NA** | **Yes** | **Yes** | **Yes** | **7** |
| **Firemark**  **1978** | **Yes** | **Yes** | **Yes** | **Yes** | **NA** | **Yes** | **Yes** | **Yes** | **7** |
| **Carmalt et al 1975** | **Yes** | **Yes** | **Yes** | **Yes** | **NA** | **Yes** | **Yes** | **NO** | **6** |
| **Singh et al 1966** | **Yes** | **Yes** | **Yes** | **Yes** | **NA** | **Yes** | **Yes** | **Yes** | **7** |
| **Hesketh 1966** | **Yes** | **Yes** | **Yes** | **Yes** | **NA** | **Yes** | **Yes** | **Yes** | **7** |
| **Cruz 1961** | **Yes** | **Yes** | **Yes** | **Yes** | **NA** | **Yes** | **Yes** | **Yes** | **7** |
|  | **Yes** | **Yes** | **Yes** | **Yes** | **NA** | **Yes** | **Yes** | **Yes** | **7** |
| **Barini 1954** | **Yes** | **Yes** | **Yes** | **Yes** | **NA** | **Yes** | **Yes** | **Yes** | **7** |

**Total= 197**

**Score 7 or more= 185**

**Score less than 7=12**

**Domains Leading explanatory questions**

Selection 1. Does the patient(s) represent(s) the whole experience of the investigator or is the selection method unclear to the extent that other patients with similar presentation may not have been reported?

Ascertainment 2. Was the exposure adequately ascertained?

3. Was the outcome adequately ascertained?

Causality

4. Were other alternative causes that may explain the observation ruled out?

5. Was there a challenge/re-challenge phenomenon?

6. Was there a dose–response effect?

7. Was follow-up long enough for outcomes to occur?

Reporting

8. Is the case(s) described with sufficient details to allow other investigators to replicate the research or to allow practitioners make

inferences related to their own practice?
