## Supplementary item-3 for "Treatment Outcome in Patients With Spinal Neurocysticercosis: A Systematic Review of Published Cases and Case Series"

**Supplementary table-3: Details of clinical features, neuroimaging characteristics and treatment outcomes in patients with spinal neurocysticercosis**

| **Reference** | **Country** | **Duration of illness** | **Clinical features** | **CSF** | **Spinal segment involvement** | **Type of spinal cord involvement** | **Imaging characteristic of lesion** | **NCC lesions elsewhere** | **Diagnostically confusing entity** | **Medical treatment** | **Surgical treatment** | **Biopsy of the lesion** | **Outcome** |
| --- | --- | --- | --- | --- | --- | --- | --- | --- | --- | --- | --- | --- | --- |
| Vijayan et al 2023 | India | 3 months | Transverse myelopathy below T4 | NA | Thoracic 3 | Intramedullary | A ring enhancing lesion | NA | Tuberculoma | NA | Lesion excision | Cysticercus granuloma | Improved able to walk |
| Tao et al 2023 | China | 3 years | Back pain  Radicular pain | NA | Lumbo-sacral region | Right L5 nerve sleeve  Subarachnoid | Cystic lesion  A large cyst and multiple small cysts | NA | Arachnoid cyst. | Albendazole | Surgical excision of all the cysts | Teania solium cyst | Improved |
| Pedrosa et al 2023 | Brazil | 3 months | Cognitive abnormalities  Difficulty in walking | Protein= 135mg/L  Cells=105 cells per μL  Glucose= low | Lumbar region | Cauda equina nerve roots clumped together | Hydrocephalus  Adhesive arachnoiditis |  | Gait apraxia  and cognitive dysfunction  Possibility of  normal pressure hydrocephalus | Albendazole with dexamethasone | NA | mNGS revealed *Taenia solium* | Improved |
| Manh et al 2023 | Viet Nam | 10 days | LMN sensory motor paraparesis | Increased protein | L4/L5 level of the lumbar spine | Intradural extramedullary | Cystic lesion with surrounding arachnoiditis | NA | NA | Albendazole with Corticosteroids | L4 and L5 decompressive laminectomy | Teania solium cyst | Improved |
| Machado et al 2023 | Brazil | 1 year | Quadriplegia | Increased protein | Cervical region | Intramedullary | Cystic lesion with scolex | An enhancing lesion in brain | NA | Albendazole with Corticosteroids | Surgical excision | Teania solium cyst | Improved |
| Lama et al 2023 | Nepal | 2 months | UMN sensory motor Paraplegia | NA | Thoracic region | Intramedullary | Cystic lesion with scolex | NA | NA | Albendazole with dexamethasone | Surgical excision | Teania solium cyst | Improved |
| Canales et al 2023 | Peru | 2 days | Back pain and paraparesis with bladder involvement | Normal | Thoracic region  D8-D9 | Intradural extramedullary | Cystic lesion with surrounding adhesive arachnoiditis | NA | NA | Albendazole with dexamethasone | Surgical excision | Teania solium cyst | Improved |
| Almeida et al 2023 | Brazil | NA | Spastic paraparesis | NA | Thoracic region  T3 T4 | Intramedullary | Cystic lesion with scolex | NA | A neoplastic lesion | NA | Surgical excision | Teania solium cyst | Improved |
| Zheng et al 2022 | China | 9 months | Spastic paraparesis | Cells and protein were markedly raised | Whole of spinal canal | Extramedullary | Multiple cystic  lesions | NA | Spinal cord tumour | Albendazole with dexamethasone | Surgical excision | Teania solium cyst | Improved |
| Yang et al 2022 | China | 1 year | Sensory motor paraparesis | NA | T11 and T12 | Intramedullary | Cystic lesion with scolex  Ring-like enhancement | NA | Cavernous malformation | NA | Surgical excision | Teania solium cyst | Improved |
|  |  | 3 years | Weakness of left leg | NA | T5 | Intramedullary | Cystic lesion with scolex | NA | Ependymoma | NA | Surgical excision | Teania solium cyst | Improved |
|  |  | 2 months | Sensory motor paraparesis | NA | T7-T8 | Intramedullary | Cystic lesion with scolex | NA | Enterogenous cyst | Albendazole for one month | Surgical excision | Teania solium cyst | Improved |
|  |  | 6 months | Sensory motor paraparesis  Back pain | NA | L1-S1 | Intramedullary | Cystic lesion with scolex | NA | Arachnoid cyst | NA | Surgical excision | Teania solium cyst | Improved |
|  |  | 6 months | Sensory motor paraparesis | NA | T8 | Intramedullary | Cystic lesion with scolex | NA | Cavernous malformation | NA | Surgical excision | Teania solium cyst | Improved |
|  |  | 3 months | Sensory motor paraparesis | NA | T7 | Intramedullary | Cystic lesion with scolex | NA | Cavernous malformation | NA | Surgical excision | Teania solium cyst | Improved |
|  |  | 3 years | Back pain | NA | T11-L1 | Extramedullary | Cystic lesion with scolex | NA | Cysticercosis | NA | Surgical excision | Teania solium cyst | Improved |
| Solanki et al 2022 | India | NA | Neck pain with radiation in arms | NA | C2-C3 level  Cervico-medullary junction to C6 | Intramedullary | Cystic lesion with scolex  Ring-like enhancement | NA | NA | Albendazole with dexamethasone | NA | NA | Improved after 6 weeks |
| Sihag et al 2022 | India | 3 months | Neck pain  Asymmetric  spastic quadriparesis | NA | C3 to C5 | Intramedullary | Cystic lesion with scolex  Ring-like enhancement | NA | NA | Albendazole with dexamethasone | Surgical excision | Teania solium cyst | Improved |
| Roy et al 2022 | India | 6 months | Severe low back pain  Cauda-equina syndrome | NA | L1-S1 | Extramedullary | Cystic lesion | NA | Epidermoid cysts  Dermoid cysts  Hydatid cysts  Tuberculomas  Intra-dural tumors  Cystic schwannoma | Albendazole with dexamethasone | Surgical excision | Teania solium cyst | Improved |
| Kus et al 2022 | USA | Sudden | Low back pain  Paraparesis | Normal | T8 | Intramedullary | Cystic lesion with scolex  Ring-like enhancement | NA | NA | Albendazole with Corticosteroids | Surgical excision | Teania solium cyst | Improved |
| Kumar et al 2022 | India | 2 years | Spastic quadriparesis | NA | C5‑C6 | Intramedullary | Cystic lesion | NA | Ependymoma | Albendazole with Corticosteroids | Surgical excision | Teania solium cyst | Improved |
| Kim et al 2022 | Korea | 10 years | Acute transverse myelitis | Increased cells | T4 | Intramedullary | A small ring-enhancing lesion with extensive cord edema from T2 to upper T6 | Multiple calcified lesions | NA | Albendazole and praziquantel with Corticosteroids | Surgical excision | Teania solium cyst | Improved |
| Gorjian and Ricks 2022 | USA | 16 years | Paraplegia | NA | L3, 4 | Intradural extramedullary | Cystic lesion | Calcified intracranial  lesions | NA | NA | Surgical excision | Teania solium cyst | Improved |
| Andino et al 2022 | USA | 3 weeks | Low back pain  Paraparesis | NA | T8 | Intramedullary | a ring-enhancing lesion | NA | Multiple sclerosis | Albendazole and praziquantel with Corticosteroids | Surgical excision | Teania solium cyst | Improved |
| Chenyu et al 2022  (in Chinese) | China | 1 year | Neck pain  Walking difficulty | NA | C4–C5 | Intradural extramedullary | Cystic lesion | Lesion in the right suprasellar cistern and gyrus rectus | NA | Repeated courses of praziquantel | Surgical excision | Teania solium cyst | Improved |
| Vadher et al 2021 | India | 2 weeks | UMN quadriparesis | NA | C7 to T2 | Intramedullary | A ring-enhancing lesion with surrounding edema | NA | NA | Albendazole with Corticosteroids | Surgical excision | Teania solium cyst | Improved |
| Rajbhandari et al 2021 | Nepal | 2 years | Low back pain | NA | L1-S1 | Extradural | Cystic lesion | NA | NA | Postoperative Corticosteroids | Surgical excision | Teania solium cyst | Improved |
| Radhakrishnan 2021 | India | 1 month | Low back pain  Cauda-equina syndrome | NA | Conus- medullaris region | Intramedullary | A ring-enhancing lesion with surrounding edema | NA | NA | Albendazole with Corticosteroids | NA | NA | Improved |
| Mediratta 2021 | India | 1 month | Neck pain | NA | C2 | Intramedullary | Cystic lesion  Ring-like enhancement  Perilesional edema from medulla to T1 | NA | Tuberculoma  Abscess,  Ependymoma  Cystic astrocytoma | Albendazole with Corticosteroids | Surgical excision | Teania solium cyst | Improved |
| Lahiri et al 2021 | India | 1 month | Low back pain  Transverse cord syndrome | Cells and protein were markedly raised | T11 | Intramedullary | A ring-enhancing lesion with surrounding edema | Multiple ring-enhancing lesion with scolex  Disseminated cysticercosis | NA | Methylprednisolone | Surgical excision of a subcutaneous nodule | Teania solium cyst | NA |
| Garg et al 2021 | India | NA | Low back pain  left lower limb radiculopathy | NA | Lumbar segments | Intradural extramedullary was operated in 2016 | Cystic lesion | NA | NA | Albendazole with Corticosteroids | Surgical excision of a subcutaneous nodule  VP Shunt  Excision of cyst in 4^th^ ventricle | Teania solium cyst | Improved initially  1 year later developed extensive spinal arachnoiditis  Vision loss due to hydrocephalus  4^th^ ventricle cyst |
| Dhar et al 2021 | India | 12 years | Low back pain  Right lower limb radiculopathy | NA | L2-3 | Intramedullary | Cystic lesion | NA | Neoplastic lesion | Albendazole with Corticosteroids | Surgical excision | Teania solium cyst | Improved |
| Chandrakanth et al 2021 | India | 1 month | Low back pain | NA | L3-L4 | Intradural  extramedullary | Cystic lesion | NA | NA | Albendazole with Corticosteroids | Surgical excision | Teania solium cyst | Improved |
| Yu et al 2020 | *Germany* | 1 month | Progressive quadriplegia | NA | C1-C2 | Intramedullary | Cystic lesion with scolex  With some enhancement | NA | NA | Albendazole with Corticosteroids | Surgical excision | Teania solium cyst | Improved |
| Walia et al 2020 | India | 1 week | Walking difficulty  Right lower limb weakness  Thoracic myelopathy T10 | NA | T7 T8 | Intramedullary | Cystic lesion with scolex  With ring enhancement | Multiple neurocysticercosis since 2008 and cognitive  dysfunction  Numerous cystic lesions | Disseminated cysticercosis | NA | Surgical excision | Teania solium cyst | Improved |
| Jobanputra et al 2020 | USA | NA | Right upper limb pain and hand numbness | NA | C5-C7 | Intramedullary | Cystic lesion with scolex  With some enhancement | NA | A primary neoplasm | NA | Surgical excision | Teania solium cyst | Improved |
| Gawande et al 2020 | India | 15 days | Seizures  Paraplegia  T7 level | NA | D8 | Intramedullary | Cystic lesion with scolex  With ring enhancement | Multi­ple small ring enhancing lesions with hole with dot appear­ance | Tuberculoma | NA | NA | NA | NA |
| Asín et al 2020 | Spain | 6 weeks | Low back pain | Increased protein | L3, displacing cauda equina roots | Multiple intradural  extramedullary cysts | Cystic lesion with scolex | Numerous  subarachnoid cysts involving basal cisterns and  communicating hydrocephalus | NA | A combination of albendazole and praziquantel  Corticosteroids | NA | NA | Improved |
| Ansari et al 2020 | India | 8 months | Low back pain  Paraplegia | NA | C7-D1 | Intramedullary | Cystic lesion | NA | NA | NA | Surgical excision | Teania solium cyst | Improved |
| Torres-Corzo et al 2019 | Mexico | 1 month | Paraparesis | NA | L4-L5 | Extramedullary | Cystic lesion | Intraventricular in past | NA | Albendazole with Corticosteroids | Surgical excision | Teania solium cyst | Improved |
|  |  | NA | Back pain | NA | L5-S1  And  Cervical region | Extramedullary  Cervical intramedullary | Cystic lesion | Intraventricular in past | NA | Albendazole with Corticosteroids | Surgical excision | Teania solium cyst | Improved |
|  |  | NA | Acute back pain | NA | L1-L4 | Subdural space | Cystic lesion | Intraventricular in past | NA | Albendazole with Corticosteroids | Surgical excision | Teania solium cyst | Improved |
| Lopez et al 2019 | Ecuador | 6 months | Progressive thoracic myelopathy T4 | NA | T4–6 and T9–11 levels | Intradural extramedullary | Multiple cystic lesions | NA | NA | Albendazole with Corticosteroids | Surgical excision | Teania solium cyst | Improved |
| Shashidhar et al 2018 | India | 2 year | Headache and seizures  2 years later  severe low backache | Increased protein and cells, low glucose  Large number of eosinophils | L2, L3 | Intradural extramedullary | Cystic lesion With ring enhancement | Had meningitis in past | Spinal hydatid cyst | Anti-helminthic drug | Surgical excision | Teania solium cyst | Improved |
| Phuyal et al 2018 | Nepal | 6 weeks | Progressive quadriparesis | NA | C3 C4 | Intramedullary | A cystic lesion with ring enhancing lesion and scolex | NA | Ependymoma | NA | Surgical excision | Teania solium cyst | Improved |
| Maste et al 2018 | India | 1 month | Low backache and right lower limb weakness | NAAT was positive for  anti‑cysticercal antibodies | T12 | Intramedullary | A cystic lesion with ring enhancing lesion | NA | Tuberculoma | Albendazole with Corticosteroids | NA | NA | Improved |
| Jeong et al 2018 (in Korean language) | Korea | 2 months | Low back pain  Paraplegia | NA | T12–L3 | Intradural extramedullary | Multiple cystic lesions with adjacent spinal arachnoiditis  Racemose cyst | Multiple cystic lesions in the brain | NA | Albendazole with Corticosteroids | Surgical excision | Teania solium cyst | Died in post-operative period |
| Agarwal et al 2018 | India | 3 weeks | Quadriparesis | NA | C3 to C4 | Intramedullary | Cystic lesion With ring enhancement | NA | NA | Praziquantel with Corticosteroids | Surgical excision | Teania solium cyst | Died in post-operative period |
| Giri et al 2018 | India | 6 months | Neck pain  Multiple cervical radiculopathies | NA | C3 to C4 | Intramedullary | Cystic lesion With ring enhancement | NA | NA | Albendazole with Corticosteroids | Surgical excision | Teania solium cyst | Improved |
| Almeida et al 2018 | Brazil | NA | Spastic paraparesis | NA | T3-T4 | Intramedullary | Cystic lesion | NA | NA | NA | Surgical excision | Teania solium cyst | NA |
| Zhang et al 2017 | China | 2 years | Back and lower limb pain  Spastic paraparesis | NA | L1/2 to S1 | Intradural extramedullary | Cystic lesion | NA | NA | NA | Surgical excision | Teania solium cyst | Improved |
| Yadav et al 2017 | India | 1 month | Progressive spastic quadriparesis | NA | C5-C6 | Intramedullary | A ring enhancing lesion | NA | NA | Albendazole with Corticosteroids | NA | NA | Improved |
| Yacoub et al 2017 | USA | 2 months | Gait difficulty  Seizures in past | NA | C4 to T4 | Intradural extramedullary | A racemose cyst extramedullary cystic lesions throughout the thoracic spine  Multi-loculated cyst in cervical region | Multiple calcified nodules | NA | Albendazole with Corticosteroids | Surgical excision | Teania solium cyst | Improved |
| Vetrivel et al 2017 | India | 7 months | Progressive spastic quadriparesis | NA | C1 to C4 | Intradural extramedullary | Cystic lesion with an enhancing wall | NA | NA | Albendazole with Corticosteroids | Surgical excision | Teania solium cyst | Improved |
| Sharma et al 2017 | India | 3 years | Low back pain | NA | L2‑S2 | Intradural extramedullary | Cystic lesion  Adhesive arachnoiditis | NA | NA | Albendazole with Corticosteroids | Surgical excision | Teania solium cyst | Improved |
| Santos et al 2017 | USA | 4 years | Low back pain | NA | L5 | Extramedullary | Several cysts were observed enveloped by nerve roots | NA | S1 radiculopathy  degenerative joint disease | NA | Surgical excision | Teania solium cyst | Improved |
| Ranjan et al 2017 | India | 1 week | Neck pain | NA | C4–C6 | Intramedullary | Cystic lesion with scolex | NA | NA | Albendazole with Corticosteroids | NA | NA | Improved |
| Pal et al 2017 | India | NA | Severe back pain  Paraparesis | NA | D9 to L5 | Intradural extramedullary | Cystic lesion with scolex | NA | Arachnoid cyst | Albendazole with Corticosteroids | Surgical excision | Teania solium cyst | Improved |
| Mesquita Filho et al 2017 | Brazil | 3 weeks | Headache and posterior neck pain  Low back pain | Increased protein and cells, low glucose | C4-C7  T12 to L1  L1  to S2 | Intradural extramedullary | Multiple cystic lesions | NA | Tuberculoma | Albendazole with Corticosteroids | Spinal biopsy | Teania solium cyst | Improved |
| Hedaoo et al 2017 | India | 3 weeks | Neck pain and progressive weakness of left upper limb | NA | C4 and C5 | Intramedullary | Cystic lesion | NA | NA | Albendazole with Corticosteroids | Spinal biopsy | Teania solium cyst | Improved |
| Hansberry et al 2017 | USA | 2 months | UMN quadriparesis | NA | CV junction to C2 | Intradural extramedullary | A large cystic lesion with peripheral enhancement  Large nodules in the lumbar region | Multiple calcific  nodules | NA | Albendazole with Corticosteroids | Surgical excision | Teania solium cyst | Improved |
| Datta et al 2017 | India | 1 month | LMN paraparesis | NA | T9 T10 | Intramedullary | Small cystic lesion with extensive perilesional edema | NA | NA | Albendazole with Corticosteroids | NA | NA | Improved  Serial MRI showed disappearance of the lesion. |
|  |  | 2 days | Low back pain  LMN paraparesis with bladder involvement | NA | T10 to T11 | Intramedullary | Multicystic lesion with scolex and heterogenous enhancement with large perilesional edema | NA | NA | Albendazole with Corticosteroids | Surgical excision | Teania solium cyst | Improved |
|  |  | 2 months | Low back pain | NA | T5 to T6 | Intramedullary | Cystic lesion with scolex and heterogenous enhancement with large perilesional edema | NA | NA | NA | NA | NA | No change in the lesion |
| Bansal et al 2017 | India | 1 year | Low back pain | NA | L5‑S1 | Intradural extramedullary | Cystic lesion | NA | Tarlov cyst | Albendazole with Corticosteroids | Surgical excision | Teania solium cyst | Improved |
| Torous and Darras 2016 | USA | 2 years | Low back pain  with acute worsening  cauda-equina syndrome | NA | L4‑S1 | Intradural extramedullary | Cystic lesion with scolex and heterogenous enhancement | NA | Malignancy of the spine | NA | Surgical excision | Teania solium cyst | Improved |
| Pant et al 2016 | India | 3 months | Spastic paraparesis | NA | T11 | Intramedullary | Cystic lesion with scolex and heterogenous enhancement | NA | Malignancy of the spine | NA | Surgical excision | Teania solium cyst | Improved |
|  |  | 1 month | Spastic paraparesis | NA | T12 to L2 | Intradural extramedullary | Cystic lesion | NA | Arachnoid cyst, epidermoid cyst, and cystic meningioma | Albendazole | Surgical excision | Teania solium cyst | Improved |
| Pratap Kumar and Ravi 2016 | India | 1 week | Cauda equina syndrome | NA | L1-S1 | Intradural extramedullary | Cystic lesion | NA | NA | Albendazole and corticosteroids | Surgical excision | Teania solium cyst | Improved |
| Mewada and Srivastava 2016 | India | 6 months | Right hemiparesis  Spastic gait | NA | C4–C5 | Intramedullary | Cystic lesion | NA | Spinal tumor | Albendazole with Corticosteroids | Surgical excision | Teania solium cyst | Improved |
| Wang and Huang 2015 | China | 4 years | Progressive vision loss, headache and thoracic back pain | Increased protein and cells, low glucose | T11  Corticomedullary junction | Perineural cysts in  the T11 foramen  Racemose cyst | Cystic lesion | Basal part of the brain | NA | Albendazole with Corticosteroids | Surgical excision | Teania solium cyst | Improved |
| Veiga et al 2015 | Portugal | 4 months | Low back pain | NA | L3-L4 to caudal end of lumbar sac | Conus terminallis | A large cystic lesion with peripheral enhancement | NA | Ependymoma | NA | Surgical excision | Teania solium cyst | Improved |
| Valsangkar et al 2015 | India | 2 months | Progressively  worsening radicular pain | NA | Conus and epiconus | Intramedullary | A large cystic lesion | NA | NA | NA | Surgical excision | Teania solium cyst | Improved |
| Salazar Noguera et al 2015 | Guatemala | 3 weeks | Paraparesis | NA | C7 to T1 | Intramedullary | A large cystic lesion | NA | Astrocytoma  and ependymoma | Albendazole | Surgical excision | Teania solium cyst | Improved |
| Ruschel et al 2015 | Brazil | 3 years | Sporadic right T5 neuropathic pain | Normal | T4-T5 | Intramedullary | A large cystic lesion with contrast enhancement | NA | NA | Corticosteroids | Surgical excision | Teania solium cyst | Improved |
| Hackius et al 2015 | Brazil | 2 weeks | Headache | Increased cells 33% eosinophils | C2 and  Cauda-equina region | Intradural extramedullary | Cystic lesions | NA | NA | Albendazole with Corticosteroids | NA | NA | Improved |
| Ganesan et al 2015 | India | 1 month | Low back pain  Cauda-equina syndrome | NA | L5–S1 | Intradural extramedullary | Cystic lesions | NA | Subarachnoid cyst or hydatid  disease | Albendazole with Corticosteroids | Surgical excision | Teania solium cyst | Improved |
| Chaurasia et al 2015 | India | 3 moths | Back pain  Spastic paraplegia | Increased protein and cells | T11 | Intramedullary | A ring-enhancing lesion with scolex and perilesional oedema | NA | NA | Albendazole with Corticosteroids | NA | NA | Improved |
| Bhardwaj 2015 | India | 1 month | Progressive quadriparesis | NA | C2 | Intramedullary | A ring-enhancing lesion with scolex and perilesional oedema | NA | NA | Albendazole with Corticosteroids | NA | NA | Improved |
| Yoo et al 2014 | Korea | NA | Low back pain | NA | L3--4 | Intradural extramedullary | Cystic lesions | NA | Arachnoid cyst | Albendazole with Corticosteroids | Surgical excision | Teania solium cyst | Improved |
| Verma et al 2014 | India | 18 months | Paraparesis with bladder dysfunction | NA | T12-L1 | Intramedullary | Cystic lesions | NA | NA | NA | Surgical excision | Teania solium cyst | Improved |
| Qazi et al 2014 | India | 6 months | Progressive spastic paraparesis | NA | D11‑L1 | Intramedullary | Cystic lesions | NA | NA | NA | Surgical excision | Teania solium cyst | Improved |
| Kim et al 2014 | Korea | 2 days | Headache and unsteady gait | NA | C7 to S1 | Intramedullary | Multiple ring-enhancing lesions | Hydrocephalus | NA | Albendazole with Corticosteroids | Surgical excision  Ventriculoperitoneal  shunting | Teania solium cyst | Improved |
| Kim et al 2014 | Korea | 1 year | Difficulty in walking | NA | T10-11 | Intramedullary | A ring-enhancing lesion | NA | Cavernous malformation | Albendazole with Corticosteroids | Surgical excision  Ventriculoperitoneal  shunting | Teania solium cyst | Improved |
| Han et al 2014 | Korea | 9 months | Sensory motor paraparesis | NA | L1 to L5 | Intradural extramedullary | A large cystic lesion | NA | NA | Albendazole with Corticosteroids | Surgical excision  Ventriculoperitoneal  shunting | Teania solium cyst | Improved |
| Ahmed and Paul 2014 | India | 3 months | Progressive spastic paraparesis | Increased protein and cells | T9 | Intramedullary | A ring-enhancing lesion with perilesional edema | NA | NA | Praziquantel with Corticosteroids | NA | NA | Improved |
| Abarrategui Yagüe et al 2014  (Article in Italian) | Spain | 3 days | Headache | Increased protein and cells and low glucose | S1-S2 | Intradural extramedullary | Cystic lesion | Multiple calcified lesions | Meningitis | Antiparasitic treatment | NA | NA | Improved |
| Iacoangeli et al 2013 | Italy | 2 years | Low back pain  Progressive paraparesis  Bladder involvement | NA | L4‑L5 | Intradural extramedullary | Cystic lesion  Racemosus type of NCC | NA | Intradural tumor of  the cauda equina | Albendazole with Corticosteroids | Surgical excision  Ventriculoperitoneal  shunting | Teania solium cyst | Improved |
| Furtado et al 2013 | India | 2 months | Progressive paraparesis | NA | T-5 | A bony vertebral lesion  The lesion extended  into the spinal canal, causing cord compression | Cystic lesion | NA | NA | Albendazole with Corticosteroids | Surgical excision  Ventriculoperitoneal  shunting | Teania solium cyst | Improved |
| De Feo et al 2013 | Italy | 1 year | Low back pain  Headache | Increased protein and cells and low glucose | D6–D8 and D10–D11 and dislocation of cauda roots | Intradural extramedullary | Cystic lesions | Multiple calcified lesions | Brucellosis  Lyme disease  Syphilis Tuberculosis | Albendazole with Corticosteroids | Surgical excision | Teania solium cyst | Improved |
| Chandramohan et al 2013 | India | 2 months | Progressive spastic paraparesis  Bladder involvement | NA | L1 | Intramedullary | Cystic lesions | NA | NA | Albendazole with Corticosteroids | NA | NA | Improved |
| Araujo et al 2013 | Brazil | 3 weeks | Progressive spastic paraparesis  Bladder involvement | NA | T2 and T3 | Intramedullary | Cystic lesions | NA | NA | Albendazole with Corticosteroids | NA | NA | Improved |
| Shin et al 2012 | Korea | 18 months | Progressive spastic paraparesis  Bladder involvement | NA | T12–S1  Whole cervical and upper thoracic involvement as well | Intradural extramedullary | Cystic lesions | NA | NA | Albendazole with Corticosteroids | Surgical excision | Teania solium cyst | Improved |
| Rice and Perera 2012 | USA | 1 month | Progressive spastic paraparesis | NA | T10 to T11 | Intramedullary | Cystic lesions | NA | NA | NA | Surgical excision | Teania solium cyst | Improved |
| Naguib et al 2012 | USA | NA | Progressive spastic paraparesis | NA | T12-L2 | Intradural extramedullary | Cystic lesions | NA | Spinal cord tumor | NA | NA | NA | NA |
| Motsepe and Ackerman 2012 | South Africa | Sudden | Severe back pain and paraparesis | NA | T12-L2 | Intradural extramedullary | Cystic lesions | NA | Spinal cord tumor | Albendazole with Corticosteroids | Surgical excision | Teania solium cyst | Improved |
| Kapu et al 2012 | India | 1 week | Severe back pain and paraparesis | NA | T12-L1 | Intradural extramedullary | Cystic lesions | NA | Inflamed epidermoid cyst | Albendazole with Corticosteroids | Surgical excision | Teania solium cyst | Improved |
| Jain et al 2012 | India | 3 years | Seizures  fever, headache and neck pain  Quadriparesis | Increased protein | C2 | Intramedullary | Cystic lesion with scolex | Ring enhancing lesions in left occipital lobe and cerebellum  Disseminated NCC | Meningitis | Albendazole with Corticosteroids | NA | NA | Improved |
| Bhowmik et al 2012 | India | 1 month | Headache | Normal | C 1 C2 | Intramedullary | Small cystic lesion | Disseminated NCC  Starry sky appearance in brain | NA | Corticosteroids only | NA | NA | Improved |
| Agale et al 2012 | India | 1 year | Paraparesis | NA | T10-T11 | Intramedullary | Cystic lesion | NA | Neurofibroma | Albendazole with Corticosteroids | Surgical excision | Teania solium cyst | Improved |
| Vij et al 2011 | India | 3 year | Severe back pain  Progressive spastic paraparesis | NA | T10-T11 | Intramedullary | Cystic lesion | NA | Ependymoma | Corticosteroids | Surgical excision | Cysticercus cyst adjacent to schwannoma | Improved |
| Seo et al 2011 | Korea | 3 months | Headache and blurred vision | Elevated protein | Conus medullaris and cauda equina | Intramedullary | Cystic lesion | NA | NA | Albendazole with Corticosteroids | Surgical excision | Teania solium cyst | Improved |
| Qi et al 2011 | China | 1 month | Progressive spastic paraparesis | NA | T4 and T5 | Intramedullary | Cystic lesion | NA | NA | NA | Surgical excision | Teania solium cyst | Improved |
| Park et al 2011 | Korea | NA | Low back pain  Progressive spastic paraparesis | NA | L5–S1 | Intradural extramedullary | Cystic lesions | NA | NA | Praziquantel with Corticosteroids | Surgical excision | Teania solium cyst | Improved |
| Lambertucci et al 2011 | Brazil | 8 months | Neck pain and upper-limb weakness | NA | C3 to C5 | Intramedullary | Cystic lesion | NA | Spinal cord tumor | Albendazole with Corticosteroids | Surgical excision | Teania solium cyst | Improved |
| Jongwutiwes et al 2011 | USA | 1 month | Progressive paraparesis | NA | L1  to L4 | Intramedullary | Cystic lesion with ring enhancing lesion | NA | Arachnoid cyst or arachnoiditis | NA | Surgical excision | Teania solium cyst | Improved |
| Heredia Mo et al 2011 | Bolivia | 3 years | Low back pain  Paraparesis | NA | L4 | Intradural extramedullary | Arachnoiditis with cyst | NA | NA | NA | Surgical excision | Teania solium cyst | Improved |
| Azfar et al 2011 | India | 3 months | Progressive spastic paraparesis | Increased protein and cells | T2 | Intramedullary | Cystic lesion with ring enhancing lesion | NA | NA | Cysticidal drug with Corticosteroids | NA | NA | Improved |
| Ahuja et al 2011 | India | 2 months | Lower limb pain | NA | L4  Filum terminale | Intradural extramedullary | Cystic lesion | NA | NA | Albendazole with Corticosteroids | NA | NA | Improved |
| Lin et al 2010 | China | 4 years | Progressive spastic quadriparesis | NA | C6, C7 | Intramedullary | Cystic lesion with scolex and ring enhancing lesion | NA | NA | Praziquantel with Corticosteroids | Surgical excision | Teania solium cyst | Improved |
| Lim et al 2010 | Korea | NA | Progressive spastic quadriparesis | NA | C2-L2 | Intradural extramedullary | Multiple cystic lesions | Normal | NA | Albendazole with Corticosteroids | Partial surgical excision | Teania solium cyst | Improved |
| Kumar et al 2010 | India | 1 month | Progressive spastic quadriparesis with bladder involvement | NA | T7 | Intramedullary | Cystic lesion with ring enhancing lesion | NA | NA | Albendazole with Corticosteroids | Surgical excision | Teania solium cyst | Improved |
| Jang et al 2010 | Korea | 3 years | Back pain and radicular  pain of the leg | NA | L4 to  S1 | Intradural extramedullary | A large cystic lesion | NA | NA | Albendazole with Corticosteroids | Surgical excision | Teania solium cyst | Improved |
| Gonçalves et al 2010 | Brazil | 9 months | Back pain and radicular  pain of the leg  Bladder involvement | Increased protein | T11 | Intramedullary | Cystic lesion with scolex | NA | NA | NA | Surgical excision | Teania solium cyst | Improved |
| Dhillon et al 2010 | Australia | 4 years | Conus medullaris syndrome | NA | T11–12 | Intramedullary | A large cystic lesion | NA | NA | NA | Surgical excision | Teania solium cyst | Improved |
| Choi et al 2010 | Korea | 2 weeks | Severe low back pain  Paraplegia | NA | L5-S1 | Intradural extramedullary | Multiple cystic lesion | NA | NA | Albendazole with Corticosteroids | Surgical excision | Teania solium cyst | Recurrent pain still persisted |
| Shin and Shin 2009 | Korea | 3 months | Paraplegia  Bladder involvement | NA | C1 to L1 | Intradural extramedullary | Multiple cysts with arachnoiditis | NA | NA | Albendazole with Corticosteroids | Surgical excision | Teania solium cyst | Improved |
| Gupta et al 2009 | India | 3 months | Backpain and difficulty in walking | Increased protein  Increased eosinophil counts | T3 and T4 | Intradural extramedullary | Multiple cysts with arachnoiditis | NA | NA | Albendazole with Corticosteroids | Surgical excision | Teania solium cyst | Improved |
| Chhiber et al 2009 | India | 15 days | Back pain  Paraplegia  Bladder involvement | NA | T4 and T7 | Intramedullary | A large cystic lesion with scolex | NA | NA | Albendazole with Corticosteroids | NA | NA | Improved |
| Mohapatra et al 2008 | India | 6 months | Headache and vision loss | Normal | Filum terminale | NA | A cystic lesion came out while shunt procedure was being done | NA | NA | Albendazole with Corticosteroids | Patient had asymptomatic spinal NCC | Teania solium cyst | Improved  Patient had asymptomatic spinal NCC |
| Kasliwal et al 2008 | India | 3 years | Headache/vomiting  Patient had hydrocephalus  Spastic quadriparesis | NA | C1-C2 | Intradural extramedullary | Cystic lesion with ring enhancing lesion | NA | NA | Albendazole with Corticosteroids | Surgical excision | Teania solium cyst | Improved |
| Izci et al 2008 | USA | 2 months | LMN paraparesis | NA | T11 to L1 | Intradural extramedullary | Cystic lesion with ring enhancing lesion | NA | NA | Albendazole with Corticosteroids | Surgical excision | Teania solium cyst | Improved |
| Edwards et al 2008 | USA | 9 months | Headache and blurry vision, papilloedema | Increased protein and cells and low glucose  Presence of eosinophils | Cervical region | Intradural extramedullary | Cystic lesion with ring enhancing lesion with arachnoiditis | Hydrocephalus | NA | Albendazole with Corticosteroids | NA | NA | Improved |
| Agrawal et al 2008 | India | 2 years | UMN paraparesis | NA | T5 T6 | Intradural extramedullary | Cystic lesion | NA | NA | Albendazole with Corticosteroids | Surgical excision | Teania solium cyst | Improved |
| Paterakis et al 2007 | Greece | 1 year | Low back pain and foot drop | NA | L5-S1 | Intradural extramedullary | Cystic lesion | NA | NA | Albendazole with Corticosteroids | Surgical excision | Teania solium cyst | Improved |
| Ahmad and Sharma 2007 | India | 2 months | Paraparesis and urinary incontinence | NA | T8 | Intramedullary | Cystic lesion with ring enhancing lesion | NA | NA | Albendazole with Corticosteroids | Surgical excision | Teania solium cyst | Improved |
|  |  | 1 month | Paraparesis and urinary incontinence | NA | T1 and T2 | Intramedullary | Cystic lesion with ring enhancing lesion and scolex | NA | NA | Albendazole with Corticosteroids | NA | NA | Improved |
| Rossi et al 2006 | Brazil | NA | Cerebellar ataxia  urinary incontinence  Sensory loss | Marked raised protein | Cervical and thoracic regions | Intradural extramedullary | Cystic lesion | cystic lesions in the subarachnoid spaces of the brain | NA | NA | Surgical excision | NA | NA |
| Guedes-Corrêa et al 2006 | Brazil | Sudden | Back pain | NA | Conus medullaris | Intramedullary | Cystic lesion with scolex | NA | NA | NA | Surgical excision | Teania solium cyst | Improved |
| Kim and Lee 2005 | Korea | Sudden | Back pain | Marked raised protein | Sacral | Intradural extramedullary | Cystic lesion | NA | NA | NA | Surgical excision | Teania solium cyst | Improved |
| Torabi et al 2004 | USA | 5 days | Intractable headache  Low back pain  UMN paraparesis | Increased protein and cells and low glucose  Presence of eosinophils | C5, T4, T5 to T9, conus meddularis, and thecal sac | Intramedullary | Enhancing lesions | Hydrocephalus Multiple intraparenchymal and intraventricular lesions | NA | Albendazole with Corticosteroids | NA | NA | Improved |
| Singh and Sahai 2004 | India | 6 months | UMN paraparesis | NA | C7-  T1 | Intramedullary | Cystic lesion | Parenchymal cysts. | NA | Albendazole with Corticosteroids | Surgical excision | Teania solium cyst | Improved |
| Jarupant et al 2004 | Thailand | 1 year | UMN paraparesis | NA | C6, and lumbar region | Intramedullary | Cystic lesion | Multiple lesions in posterior fossa | NA | Albendazole with Corticosteroids | Surgical excision | Teania solium cyst | Died |
| Delobe et al 2004 | French Guiana | 2 months | Cauda equina syndrome | Increased protein and cells and low glucose  Presence of eosinophils (Eosinophils 57%) | L3–L4 level, compressing cauda equina | Intradural extramedullary | Cystic lesion | Two focal cortical enhancing lesions | NA | Albendazole with Corticosteroids | Surgical excision | Teania solium cyst | Improved |
| Jang et al 2003  (Article is in Korean language) | Korea | 1 year | Upper limb weakness | Increased protein and cells | C4-5 | Intramedullary | Cystic lesion with peripheral enhancement | NA | NA | Albendazole with Corticosteroids | Surgical excision | Teania solium cyst | Improved |
| Costa Junior et al 2003 | Brazil | 6 months | Low back pain  Paraparesis | NA | L1-S1 in cauda equina region | Intradural extramedullary | Multiple cystic lesions  Racemose cysts | NA | NA | NA | Surgical excision | Teania solium cyst | Improved |
| Sheehan et al 2002 | USA | 4 months | Had accident was screened for neck injury | NA | A lesion at C1–2 | Intramedullary | Cystic | NA | Neck injury | Praziquantel with Corticosteroids | Surgical excision | Teania solium cyst | Improved |
| Yoon et al 2002 (Article is in Korean language) | Korea | 1 month | Quadriplegia  Seizures in past | NA | Upper cervical region | Intradural extramedullary | Multiple cystic lesions  Racemose cysts | Multiple cystic lesions in posterior fossa  Racemose cysts | NA | Albendazole with Corticosteroids | Surgical excision | Teania solium cyst | Improved |
| Muzumdar et al 2002 | India | 2 months | Spastic paraparesis | NA | T7 and T8 | Intramedullary | Cystic lesion with peripheral enhancement  Scolex | NA | NA | Albendazole with Corticosteroids | Surgical excision | Teania solium cyst | Improved |
|  |  | 3 days | Spastic paraparesis | NA | T8 and T9 | Intramedullary | Cystic lesion with peripheral enhancement  Scolex | NA | NA | Albendazole with Corticosteroids | Surgical excision | Teania solium cyst | Improved |
| Alsina et al 2002 | USA | 1 month | Progressive spastic paraparesis | NA | L2 L3 | Intradural extramedullary | Large cyst | Had brain lesions in the past | NA | Praziquantel with Corticosteroids | Surgical excision | Teania solium cyst | Improved |
|  |  | 2 weeks | Progressive quadriparesis | NA | C-5 to T-1 | Intradural extramedullary | Large cyst | NA | NA | Corticosteroids | Surgical excision | Teania solium cyst | Improved |
|  |  | 1 week | Progressive quadriparesis | NA | C-5 | Intradural extramedullary | Large cyst | NA | NA | Corticosteroids | Surgical excision | Teania solium cyst | Improved |
|  |  | NA | Headache | NA | Foramen magnum | Intradural extramedullary | Large cyst | NA | NA | Praziquantel with Corticosteroids | Surgical excision | Teania solium cyst | Headache persisted |
|  |  | NA | Progressive spastic quadriparesis | NA | C1 | Intramedullary | Cystic lesion | NA | NA | Corticosteroids | NA | NA | Improved |
|  |  | NA | Difficulty in walking | NA | T7–9 | Intradural extramedullary | Cystic lesion | NA | NA | Praziquantel with Corticosteroids | Surgical excision | Teania solium cyst | Improved |
| Parmar et al 2001 | India | NA | Progressive paraparesis | NA | T8 T9 | Intramedullary | Cystic lesion with scolex | NA | NA | Albendazole with Corticosteroids | Surgical excision | Teania solium cyst | Improved |
|  |  | 2 months | Progressive paraparesis | NA | T7 T8 | Intramedullary | Cystic lesion with scolex | NA | NA | Albendazole with Corticosteroids | Surgical excision | Teania solium cyst | Improved |
|  |  | 1 month | Pain in neck | NA | C5 | Intramedullary | Cystic lesion with scolex | NA | NA | Albendazole with Corticosteroids | NA | NA | Improved |
|  |  | NA | Progressive quadriparesis | NA | C4 C5 | Intramedullary | Cystic lesion with scolex | NA | NA | Albendazole with Corticosteroids | NA | NA | Improved |
|  |  | NA | Progressive paraparesis | NA | T8 T9 | Intramedullary | Cystic lesion with scolex | Disseminated NCC | NA | Albendazole with Corticosteroids | NA | NA | Improved |
|  |  | 3 months | Progressive paraparesis | NA | T10 T11 | Intramedullary | Cystic lesion with scolex | Brain lesions were present | NA | Albendazole with Corticosteroids | NA | NA | Improved |
| Mathuriya et al 2001 | India | 10 weeks | Back pain and paraparesis | Increased protein and cells | T1 | Intramedullary | Cystic lesion with scolex | NA | NA | NA | Surgical excision | Teania solium cyst | Improved |
|  |  | 7 months | Spastic paraparesis | NA | T2 | Intramedullary | Cystic lesion with scolex | NA | NA | NA | Surgical excision | Teania solium cyst | Improved |
|  |  | 2 years | Spastic paraparesis | NA | T11 | Intramedullary | Cystic lesion with scolex | NA | NA | NA | Surgical excision | Teania solium cyst | Improved |
| Homans et al 2001 | USA | 2 weeks | Back pain | NA | T11 and T12 | Intramedullary | Cystic lesion with scolex | NA | Ependymoma | NA | Surgical excision | Teania solium cyst | Improved |
| Sahoo 2000 | India | 6 months | Neck pain and upper limb weakness | NA | C 5 | Intramedullary | Cystic lesion with scolex | NA | NA | Albendazole with Corticosteroids | Surgical excision | Teania solium cyst | Improved |
| Gaur et al 2000 | India | 2 months | Spastic paraparesis | Normal | T9 | Intramedullary | Cystic lesion with scolex | NA | NA | Albendazole with Corticosteroids | NA | NA | Improved |
|  |  | 5 months | Spastic paraparesis | Normal | T5 T6 | Intramedullary | Cystic lesion with scolex | NA | NA | Albendazole with Corticosteroids | NA | NA | Improved |
| Dantas et al 1999  (Article is in Portuguese  language) | Brazil | NA | Progressive quadriparesis | NA | C4 C5 | Intramedullary | Cystic lesion with scolex | NA | NA | NA | Surgical excision | Teania solium cyst | Improved |
| Ciftci et al 1999 | USA | 2 months | Neck pain | NA | C2 | Intradural extramedullary | Cystic lesion | Basal  cistern, cisterna magna, and cervical subarachnoid space | NA | NA | NA | NA | NA |
| Rosahl and Samii 1998  Article in German | Germany | NA | Quadriparesis | NA | Cervical region | Intramedullary | Cystic lesion with scolex | Basal  cistern, cisterna magna, and cervical subarachnoid space | NA | NA | NA | NA | NA |
| Mohanty et al 1998 | India | 5 months | Quadriparesis | NA | C4 C5 C6  On myelography | Intradural extramedullary | Cystic lesion | NA | NA | NA | Surgical excision | Teania solium cyst | Improved |
| Lau et al 1998 | Hong Kong | 1 year | Paraparesis  right sensorineural  deafness | NA | Lumbosacral subarachnoid space | Intramedullary | Ill-defined | Multilobulated intra- and extra-axial lesions | Metastasis | NA | Surgical excision | Teania solium cyst | NA |
| Garg and Nag 1998 | India | 2 days | Paraparesis | Increased protein and cells | T9 | Intramedullary | Cystic lesion with scolex | NA | NA | Albendazole with Corticosteroids | NA | NA | Improved |
|  |  | 4 days | Paraparesis | Increased protein and cells | T8 | Intramedullary | Cystic lesion with scolex | Subcutaneous cysticercal cyst | NA | Albendazole with Corticosteroids | NA | NA | Improved |
| Escobar et al 1998 | Mexico | 1 month | Quadriparesis | NA | C5-C6 | Intramedullary | Cystic lesion with scolex | NA | NA | NA | Surgical excision | Teania solium cyst | NA |
| Davies et al 1996 | Australia | 3 years | Progressive quadriparesis | NA | Cervical and CV junction | Intradural extramedullary | Multilocular cystic collection | Patient had shunt surgery | Arachnoid cyst | Praziquantel with Corticosteroids | Surgical excision | Teania solium cyst | Improved |
| Corral et al 1996 | Mexico | NA | Seizures  Progressive quadriparesis | NA | Cervical region | Intramedullary | Cystic lesion | Multiple cystic lesions of the brain | NA | Repeated courses of albendazole and praziquantel with Corticosteroids | NA | NA | Improved |
| Prasathapong 1995 | Thailand | 2 weeks | Progressive paraparesis | NA | T10 T11 | Intramedullary | Cystic lesion | NA | NA | NA | Surgical excision | Teania solium cyst | Improved |
| Kim et al 1995 | Korea | 6 months | Upper limb weakness | NA | C2 | Intramedullary | Cystic lesion | NA | NA | NA | Surgical excision | Teania solium cyst | Improved |
|  |  | 8 months | Back pain | NA | L1 to L5-S1 | Intradural extramedullary | Cystic lesion | Multiple cystic lesions of the brain | NA | NA | NA | NA | NA |
|  |  | 6 months | Upper limb weakness  Diplopia | NA | C1 C2 | Intramedullary | Cystic lesion | Multiple cystic lesions of the brain | NA | Praziquantel | Surgical excision | Teania solium cyst | Improved |
|  |  | NA | Upper limb weakness  Following praziquantel therapy | NA | C1-C6 | Intradural extramedullary | Cystic lesion | Multiple cystic lesions of the brain | NA | Praziquantel | NA | NA | Improved |
| Gallani et al 1992  “Article is in Portuguese” | Brazil | 6 months | Low back pain | NA | Thoracolumbar myelography  T11 and T12 | Intramedullary | Cystic lesion | NA | NA | NA | Surgical excision | Teania solium cyst | NA |
|  |  | 2 months | Progressive paraparesis | Normal | L1 and L2 | Intradural extramedullary | Cystic lesion | NA | NA | NA | Surgical excision | Teania solium cyst | Improved |
| Bandres et al 1992 | USA | 2 months | Neck pain and low back pain  Later unconsciousness | Increased protein and cells | C1 to C3  L3 | Intradural extramedullary | Cystic lesion | Hydrocephalus | NA | Repeated courses of praziquantel and albendazole with corticosteroids | NA | NA | Improved |
| Palasis and Drevelengas 1991 | Greece | 4 months | Back pain | NA | Myelogram showed filling defects in cauda equina | NA | Cystic lesion | NA | NA | NA | Surgical excision | Teania solium cyst | Improved |
| Venkataramana et al 1989 | India | 6 months | Progressive paraparesis | Increased protein | T2 | Myelogram showed  intradural extramedullary  lesion | Cystic lesion | NA | Tuberculosis | NA | Surgical excision | Teania solium cyst | Improved |
|  |  | 4 months | Progressive quadriparesis | Increased protein | C3 | Myelogram showed  Intramedullary lesion | Cystic lesion | NA | NA | NA | Surgical excision | Teania solium cyst | Improved |
| Sperlescu et al 1989  “Article is in Portuguese” | Brazil | NA | Progressive paraparesis | Increased protein and cells | The conus medullaris and cauda equina | Intradural extramedullary | Cystic lesion | NA | NA | NA | Surgical excision | Teania solium cyst | Improved |
|  |  | 1 year | Low back pain | Increased cells | the cauda equina | Intradural extramedullary | Cystic lesion | NA | NA | NA | Surgical excision | Teania solium cyst | Improved |
| Vlok and Wells 1988 | South Africa | 1 year | Progressive paraparesis | NA | Destruction of T11 vertebrae | Bony lesion | Grape like cystic lesion | NA | NA | NA | Surgical excision | Teania solium cyst | Improved |
| Castillo et al 1988 | USA | 1 week | Limb weakness and limb pain | Normal | C3 to C7, T3 to TS and at the conus medullaris | Intramedullary | Cystic lesion | NA | NA | NA | Surgical excision | Teania solium cyst | Improved |
| Sharma et al 1987 | India | 3 months | Progressive paraparesis | Increased protein and cells | T5 on  Myelogram | Intramedullary | Cystic lesion | NA | NA | NA | Surgical excision | Teania solium cyst | Improved |
| Savoiardo et al 1986 | Italy | 2 months | Progressive paraparesis | Increased protein and cells | T11, T12, L1-3  Conus medullaris  on  Myelogram | Intradural extramedullary | Cystic lesion | X-ray films of the thighs which showed  shaped calcifications | NA | Praziquantel with Corticosteroids | NA | NA | Improved |
| Holtzman et al 1986 | USA | NA | Progressive paraparesis | Increased protein | T4 | Intramedullary | Cystic lesion | NA | NA | NA | Surgical excision | Teania solium cyst | Improved |
| Kim et al 1985 | USA | 2 years | Headache and vision loss | Increased protein and cells and low glucose | L3 | Intradural extramedullary | Cystic lesion | Hydrocephalus and shunt was placed  Basal cysts | NA | NA | NA | NA | NA |
| McDonald et al 1979 | USA | 1 month | Flaccid Paraparesis | NA | Cauda equina region  By Myelography | Intradural extramedullary | Racemose cysticercosis  Diffuse arachnoiditis | NA | NA | Prednisolones | Surgical excision | Teania solium cyst | Improved |
| Akiguchi et al 1979 | Japan | 3 years | Back pain  Cauda-equina syndrome | Markedly raised protein | T12  By Myelography | Intramedullary | Cystic lesion | NA | NA | NA | Surgical excision | Teania solium cyst | Improved |
| Firemark  1978 | USA Immigrant | 5 years | Back pain  Progressive paraparesis | Normal | L3 L4  By Myelography | Intradural extramedullary | Cystic lesion  Diffuse arachnoiditis | NA | NA | NA | Surgical excision | Teania solium cyst | Improved |
| Carmalt et al 1975 | USA | NA | Headache and back pain  Altered sensorium | Increased protein and cells | Multiple filling defects in lumbo-thoracic region on myelography | Multiple subarachnoid cysts | NA | NA | NA | NA | Surgical excision | Teania solium cyst | NA |
| Singh et al 1966 | India | 10 days | Paraparesis  Seizures | Increased protein | T8  On myelography | NA | Cystic lesion | Muscle calcifications | NA | NA | Surgical excision | Teania solium cyst | Improved |
| Hesketh 1966 | Singapore | 1 week | Back pain | Increased protein and cells | Multiple filling defects in thoracic region on myelography | NA | Cystic lesion | NA | Spinal cord tumor | NA | Surgical excision | Teania solium cyst | Improved |
| Cruz 1961 | Brazil | 6 months | Back pain  Paraparesis | Increased protein and cells | Myelography= lesion at T12  Cauda equina | NA | Cystic lesion  Arachnoiditis | NA | NA | NA | Surgical excision | Teania solium cyst | Improved |
|  |  | 18 months | Back pain  Paraparesis | Increased cells | Myelography= lesion at T8 | NA | Cystic lesion  Arachnoiditis | NA | Corticosteroids | NA | Surgical excision | Teania solium cyst | Improved |
| Barini 1954 | Brazil | 4 months | Back pain  Paraparesis | Increased cells | Myelography= T10 and T11 | Intramedullary | Cystic lesion  Arachnoiditis | NA | NA | NA | Surgical excision | Teania solium cyst | Improved |

**References in a chronological order**

1. Vijayan SP, Gerber C, Basu A, Mhatre R. Isolated spinal intramedullary neurocysticercosis: Case report and review of literature. Indian Spine Journal. 2023;6(1):96-100.
2. Tao B, Li T, Ji K, Shang A. Spinal nerve root sleeve cysticercosis: a case report and review of the literature. J Med Case Rep. 2023 Feb 22;17(1):80. doi: 10.1186/s13256-022-03733-9.
3. Pedrosa DA, Bruniera Peres Fernandes G, Filipe de Souza Godoy L, C Felício A, Pinho JR, Mário Doi A, de Araújo Gleizer R. Neurocysticercosis: diagnosis via metagenomic next-generation sequencing. Pract Neurol. 2023 Nov 23;23(6):509-511.
4. Manh BH, Dat T, Hai VT, He DV, Ha DD, Que NV, Duc NM. Spinal cysticercosis: A case report. Radiol Case Rep. 2023 Jul 6;18(9):3269-3273. doi: 10.1016/j.radcr.2023.06.037.
5. Machado S, Ewaldo Lindorfer Neto E, de Carvalho Dornelas B, de Martino Luppi A, Henrique de Oliveira E, Cesar Marinho Dias P, Dos Reis MQ, Dos Santos DF. Intramedullary Neurocysticercosis: A Case Report. Neurology. 2023 Nov 27;101(22):1023-1024. doi: 10.1212/WNL.0000000000207911.
6. Lama SM, Paudel K, Shalikhe N, Yadav PK, Lamichhane S. Isolated spinal Intramedullary Neurocysticercosis: A rare case report and review of literature. Nepal Journal of Neuroscience. 2023;20(3):66-9.
7. Canales D, Araujo-Chumacero MM, Vences MA, Latorre A. Acute dorsal myelopathy as an atypical presentation of spinal neurocysticercosis. Revista Chilena de Infectologia. 2023;40(1):66-9.
8. Almeida C Jr, de Almeida GC, Pentiado JAM Jr, Konichi Dias R. Teaching NeuroImages: Spinal intramedullary cysticercosis: The pseudotumoral form. Neurology. 2018 Sep 18;91(12):e1202-e1203. doi: 10.1212/WNL.0000000000006206.
9. Zheng X, Wang F, Wang L, Li X, Li J, Huang M, Zou Y. A Rare Case of Cysticercosis Involving the Whole Spinal Canal. Acta Parasitol. 2022 Mar;67(1):569-572. doi: 10.1007/s11686-021-00486-1.
10. Yang C, Liu T, Wu J, Xie J, Yu T, Jia W, Yang J, Xu Y. Spinal cysticercosis: a rare cause of myelopathy. BMC Neurol. 2022 Feb 22;22(1):63. doi: 10.1186/s12883-022-02589-2.
11. Solanki M, Reddy Gayam VR, Agrawal K, Jindal N, Yadav K. Isolated Intramedullary Cervical Spinal Cord Cysticercosis- A Case Report. Journal of Clinical and Diagnostic Research. 2022;16(5).
12. Sihag RK, Pannem R, Khanderia RA, Arora RK. Cysticercosis Presenting as an Isolated Cervical Intramedullary Lesion: A Rare Benign Condition at a Dangerous Location. Indian Journal of Neurosurgery. 2022 Aug;11(02):185-7.
13. Roy SS, Barman A, Viswanath A, Sahoo J. Isolated neurocysticercosis of the spine presenting with low back pain and cauda equina syndrome: a case report. Spinal Cord Ser Cases. 2022 Jul 26;8(1):70. doi: 10.1038/s41394-022-00535-5.
14. Kus J, Panah E, Rosenblum J, Bashir M. Isolated Spinal Cord Neurocysticercosis. J Radiol Case Rep. 2022 Oct 1;16(10):1-7. doi: 10.3941/jrcr.v16i10.4543.
15. Kumar A, Bhaisora KS, Sasapardhi S, Srivastava AK. Cervical Intramedullary Cysticercosis. Journal of Spinal Surgery. 2022 Apr 1;9(2):134-5.
16. Kim HJ, Kim SH, Jeong HS, Kim BJ. Intramedullary parasite eggs, latent for three decades, mimicking acute transverse myelitis. BMC Infect Dis. 2022 Jan 4;22(1):9. doi: 10.1186/s12879-021-07013-7.
17. Gorjian M, Ricks C. Late Onset Symptomatic Enlargement of Treated Spinal Intradural Neurocysticercosis Cyst. Neurohospitalist. 2022 Oct;12(4):708-710. doi: 10.1177/19418744221108304.
18. Andino D, Tsiang JT, Pecoraro NC, Jani R, Iordanou JC, Zakaria J, Borys E, Pasquale DD, Nockels RP, Schneck MJ. Case report and review of literature: Isolated intramedullary spinal neurocysticercosis. Front Neurol. 2022 Nov 10;13:1030468. doi: 10.3389/fneur.2022.1030468.
19. Chenyu L, Yuanhao L, Shaoxiong W, Shuang Z, Changkai M, Xiukun L, Yunqian L. A case of hydrocephalus caused by cerebral cysticercosis combined with cervical subdural extramedullary. Chinese Journal of Neurosurgery. 2022;38(2):197-8. DOI: 10.3760/cma.j.cn112050-20210624-00308.
20. Vadher A, Raval MR, Shah SD, Patel KG, Sharma K. A Rare Case of Isolated Intramedullary Spinal Cord Cysticercosis. Cureus. 2021 May 6;13(5):e14864. doi: 10.7759/cureus.14864.
21. Rajbhandari S, Gurung P, Yadav J, Rajbhandari P, Acharya S, Pant B. A case report of multiple isolated intradural neurocysticercosis of the lumbo-sacral spine. Int J Surg Case Rep. 2021 Oct;87:106434. doi: 10.1016/j.ijscr.2021.106434.
22. Radhakrishnan DM, Bhasi A, Saurya S, Shree R, Kumar N. Spinal Intramedullary Neurocysticercosis An Unusual Cause of Paraparesis. J Assoc Physicians India. 2021 Sep;69(9):11-12.
23. Mediratta S. A rare case of primary high cervical intramedullary cysticercosis: Uncomplicated surgery but a preoperative diagnostic predicament. Indian Spine Journal. 2021;4(2):255-9.
24. Lahiri D, Chowdhury A, Dubey S, Ray BK. Acute dorsal myelopathy resulting from intramedullary cysticercus: a case report. J Med Case Rep. 2021 Mar 17;15(1):139. doi: 10.1186/s13256-021-02693-w.
25. Garg K, Vij V, Garg A, Singh M, Chandra PS. "Malignant" Craniospinal Neurocysticercosis: A Rare Case. World Neurosurg. 2021 Feb;146:95-102. doi: 10.1016/j.wneu.2020.10.121.
26. Dhar A, Dua S, Singh H. Isolated Intramedullary Lumbar Spine Neurocysticercosis: A Rare Occurrence and Review of Literature. Surg J (N Y). 2021 Dec 15;7(4):e327-e336. doi: 10.1055/s-0041-1739118.
27. Chandrakanth K, Reddy RS, Multani KM. Lumbar subarachnoid neurocysticercosis: a case report with literature review. Indian Journal of Neurosurgery. 2021;11(03):271-3. DOI https://doi.org/ 10.1055/s-0041-1722833
28. Yu Y, Jin Z, Ma H, Chen F. Spinal intramedullary cysticercosis with syringomyelia: a case report. Int J Clin Exp Pathol. 2020 Oct 1;13(10):2593-2598.
29. Walia GK, Baveja P, Verma S, Kumar S, Jain A, Dudhal R. Intramedullary Cysticercosis-An Uncommon Entity Even in the Presence of Intracranial Neurocysticercosis. Annals of Pathology and Laboratory Medicine. 2020;7(9). DOI: 10.21276/APALM.2900.
30. Jobanputra K, Raj K, Yu F, Agarwal A. Intramedullary Neurocysticercosis Mimicking Cord Tumor. J Clin Imaging Sci. 2020 Feb 28;10:7. doi: 10.25259/JCIS_165_2019.
31. Gawande V, Saoji K, Nair A, Saoji K. Radiological findings of spinal neurocysticercosis. International Journal of Current Research and Review. 2020;12(18):164-8. DOI: 10.31782/IJCRR.2020.121824.
32. Pérez-Jacoiste Asín MA, Calleja-Castaño P, Hilario A. Lumbosacral Radiculopathy as the Clinical Presentation of Neurocysticercosis. Am J Trop Med Hyg. 2020 Jun;102(6):1166-1167. doi: 10.4269/ajtmh.19-0757.
33. Ansari A, Agrawal S, Riyaz S. Spinal intramedullary cysticercosis mimicking spinal tumour. Romanian Neurosurgery. 2020:89-91. DOI: 10.33962/roneuro-2020-011.
34. Torres-Corzo JG, Islas-Aguilar MA, Cervantes DS, Chalita-Williams JC. The Role of Flexible Neuroendoscopy in Spinal Neurocysticercosis: Technical Note and Report of 3 Cases. World Neurosurg. 2019 Oct;130:77-83. doi: 10.1016/j.wneu.2019.06.194.
35. Lopez S, Santillan F, Diaz JJ, Mogrovejo P. Spinal cord compression by multiple cysticercosis. Surg Neurol Int. 2019 Jun 7;10:94. doi: 10.25259/SNI-46-2019.
36. Shashidhar A, Savardekar AR, Mundlamuri RC, Netravathi M, Nalini A, Chickabasaviah YT, Arivazhagan A, Rao MB. Chronic eosinophilic meningitis as a manifestation of isolated spinal neurocysticercosis: A rare case and a review of literature. Neurol India. 2018 Mar-Apr;66(2):561-564. doi: 10.4103/0028-3886.227297.
37. Phuyal S, Rajbhandari B, Sedain G, Shilpakar SK. An unusual case of Cervical intramedullary neurocysticercosis mimicking ependymoma. Nepal Journal of Neuroscience. 2018;15(3):62-5.
38. Maste PS, Lokanath YK, Mahantshetti SS, Soumya S. Isolated Intramedullary Spinal Cysticercosis: A Case Report with Review of Literature of a Rare Presentation. Asian J Neurosurg. 2018 Jan-Mar;13(1):154-156. doi: 10.4103/1793-5482.180894.
39. Jeong Y-H, Lee Y-S, Eun D-C, Byun C-W. Intradural extramedullary cysticercosis involving the thoracolumbar spinal canal in a patient with cerebral cysticercosis. Journal of the Korean Orthopaedic Association. 2018;53(4):369-73. https://doi.org/10.4055/jkoa.2018.53.4.369.
40. Agarwal A, Bhatia R, Sharma BS. Rare case of intramedullary spinal cysticercosis-a case report. Journal of Evidence Based Medicine and Healthcare 2018 5(52), 3611-3613. DOI: 10.18410/jebmh/2018/736.
41. Giri SA, Diyora B, Giri D, Giri P, Sharma A. A rare case of isolated cervical intramedullary cysticercosis: A surgical dilemma. Journal of Spinal Surgery. 2018;5(2):82. 10.5005/jp-journals-10039-1176.
42. Almeida C Jr, de Almeida GC, Pentiado JAM Jr, Konichi Dias R. Teaching NeuroImages: Spinal intramedullary cysticercosis: The pseudotumoral form. Neurology. 2018 Sep 18;91(12):e1202-e1203. doi: 10.1212/WNL.0000000000006206.
43. Zhang S, Hu Y, Li Z, Zhao L, Wang Z. Lumbar spinal intradural neurocysticercosis: A case report. Experimental and Therapeutic Medicine. 2017;13(6):3591-3. DOI: 10.3892/etm.2017.4403.
44. Yadav K, Garg D, Kaushik JS, Vaswani ND, Dubey R, Agarwal S. Intramedullary Neurocysticercosis Successfully Treated with Medical Therapy. Indian Journal of Pediatrics. 2017;84(9):725-6. DOI: 10.1007/s12098-017-2353-x.
45. Yacoub HA, Goldstein I, El-Ghanem M, Sharer L, Souayah N. Spinal racemose cysticercosis: case report and review. Hospital Practice. 2017;45(3):99-103. DOI: 10.1080/21548331.2017.1325704.
46. Muralidharan V, Nair BR, Patel B, Rajshekhar V. Primary Intradural Extramedullary Cervical Spinal Cysticercosis. World Neurosurg. 2017 Oct;106:1052.e5-1052.e11. doi: 10.1016/j.wneu.2017.07.008.
47. Sharma R, Garg K, Agarwal D, Garg A, Sharma MC, Sharma BS, Mahapatra AK. Isolated primary intradural extramedullary spinal cysticercosis. Neurol India. 2017 Jul-Aug;65(4):882-884. doi: 10.4103/neuroindia.NI_98_17.
48. Santos PJ, Suzuki S, Vadera S. Back Pain and Spinal Cysticercosis. J Clin Neurol. 2017 Jan;13(1):114-115. doi: 10.3988/jcn.2017.13.1.114. Epub 2016 Nov 17.
49. Ranjan R, Tulika, Chand S, Agnihotri A. Solitary Intramedullary Cervical Cysticercosis without Neurological Deficit: A Rare Case Report. J Pediatr Neurosci. 2017 Jan-Mar;12(1):99-101. doi: 10.4103/jpn.JPN_162_16.
50. Pal A, Biswas C, Ghosh TR, Deb P. A rare case of recurrence of primary spinal neurocysticercosis mimicking an arachnoid cyst. Asian J Neurosurg. 2017;12(2):250-2. DOI: 10.4103/1793-5482.144176.
51. Mesquita Filho PM, Azambuja Junior ND, Vanzin JR, Annes RD, Varela DL, Araújo MAD, et al. Spinal neurocysticercosis: A rare variant of a common parasitic infection. Brazilian Neurosurgery. 2017;36(1):66-70. DOI: 10.1055/s-0035-1571268.
52. Hedaoo K, Garg S, Thanvi S, Agay AK, Nagocha V, Rao M. Cervical spine intramedullary cysticercosis in a young adult-a case report and literature review. International Journal of Research in Medical Sciences. 2017;5(5):2260. DOI: 10.18203/2320-6012.ijrms20171884.
53. Hansberry DR, Agarwal N, Sharer LR, Goldstein IM. Minimally manipulative extraction of polycystic cervical neurocysticercosis. Eur Spine J. 2017 May;26(Suppl 1):63-68. doi: 10.1007/s00586-016-4763-2.
54. Datta SGS, Mehta R, Macha S, Tripathi S. Primary Spinal Intramedullary Neurocysticercosis: A Report of 3 Cases. World Neurosurg. 2017 Sep;105:1037.e1-1037.e7. doi: 10.1016/j.wneu.2017.05.168.
55. Bansal S, Suri A, Sharma MC, Kakkar A. Isolated lumbar intradural extra medullary spinal cysticercosis simulating tarlov cyst. Asian J Neurosurg. 2017 Apr-Jun;12(2):279-282. doi: 10.4103/1793-5482.150225.
56. Torous VF, Darras N. A lumbar canal cystic mass lesion in a man with a history of chronic lower back pain. Neuropathology. 2016 Feb;36(1):103-6. doi: 10.1111/neup.12228.
57. Pant I, Chaturvedi S, Singh G, Gupta S, Kumari R. Spinal cysticercosis: A report of two cases with review of literature. J Craniovertebr Junction Spine. 2016 Oct-Dec;7(4):285-288. doi: 10.4103/0974-8237.193261.
58. Kumar NP, Ravi K. Isolated cauda equina cysticercosis-a rare cause of cauda equina syndrome. Journal of Evidence Based Medicine and Healthcare. 2016; 3(72), 3945-3948. DOI: 10.18410/jebmh/2016/842.
59. Mewada T, Srivastava AK. Isolated cervical intramedullary cysticercosis. Neurol India. 2016 Jan-Feb;64(1):188-9. doi: 10.4103/0028-3886.173665.
60. Wang DD, Huang MC. Cervicomedullary neurocysticercosis causing obstructive hydrocephalus. J Clin Neurosci. 2015 Sep;22(9):1525-8. doi: 10.1016/j.jocn.2015.03.031.
61. Veiga A, Matas A, Gabriel J, Martins M. Conus terminallis neurocysticercosis: a rare cause of lumbar radiculopathy. J Neurol Neurophysiol. 2015;6(265.10):4172. DOI: 10.4172/2155-9562.1000265.
62. Valsangkar SA, Kharosekar HU, Palande DA, Velho V. Isolated conus-epiconus neurocysticercosis. Neurol India. 2015 Jan-Feb;63(1):119-20. doi: 10.4103/0028-3886.152686.
63. Salazar Noguera EM, Pineda Sic R, Escoto Solis F. Intramedullary spinal cord neurocysticercosis presenting as Brown-Séquard syndrome. BMC Neurol. 2015 Jan 16;15:1. doi: 10.1186/s12883-014-0245-5.
64. Ruschel LG, Merida KB, Agnoletto GJ, de Souza Machado GA, Navarrete FAC, Ramina R. Surgical Resection of Isolated Intramedullary Neurocysticercosis. JBNC-Journal Brasileiro de Neurocirurgia. 2015;26(3):223-6.
65. Hackius M, Pangalu A, Semmler A. Neurological picture. Isolated spinal neurocysticercosis. J Neurol Neurosurg Psychiatry. 2015 Feb;86(2):234-5. doi: 10.1136/jnnp-2013-307142.
66. Ganesan S, Acharya S, Kalra KL, Chahal R. Intradural Neurocysticercosis of Lumbar Spine: A Case Report. Global Spine J. 2015 Aug;5(4):e1-4. doi: 10.1055/s-0034-1394125.
67. Chaurasia RN, Mishra VN, Jaiswal S. Spinal cysticercosis: an unusual presentation. BMJ Case Rep. 2015 Jan 23;2015:bcr2014207966. doi: 10.1136/bcr-2014-207966.
68. Bhardwaj N. Spinal Intramedullary Cysticercosis: A Rare Diagnostic Dilemma. J Emerg Med. 2015 Sep;49(3):e79-80. doi: 10.1016/j.jemermed.2014.12.094.
69. Yoo M, Lee CH, Kim KJ, Kim HJ. A case of intradural-extramedullary form of primary spinal cysticercosis misdiagnosed as an arachnoid cyst. J Korean Neurosurg Soc. 2014 Apr;55(4):226-9. doi: 10.3340/jkns.2014.55.4.226.
70. Verma SK, Gupta VK, Swamy MN, Pathak HC. Cysticercosis of conus medullaris: a case report and literature review. Indian Journal of Neurosurgery. 2014;3(01):054-6. DOI:10.4103/2277-9167.132010.
71. Qazi Z, Ojha BK, Chandra A, Singh SK, Srivastava C, Patil TB. Isolated intramedullary spinal cord cysticercosis. J Neurosci Rural Pract. 2014 Nov;5(Suppl 1):S66-8. doi: 10.4103/0976-3147.145209.
72. Kim SW, Wang HS, Ju CI, Kim DM. Acute hydrocephalus caused by intraspinal neurocysticercosis: case report. BMC Res Notes. 2014 Jan 2;7:2. doi: 10.1186/1756-0500-7-2.
73. Kim M, Rhim SC, Khang SK. Intramedullary spinal cysticercosis: a case report and review of literature. Korean J Spine. 2014 Jun;11(2):81-4. doi: 10.14245/kjs.2014.11.2.81.
74. Han SB, Kwon HJ, Choi SW, Koh HS, Kim SH, Song SH, Youm JY. Lumbar intradural neurocysticercosis: a case report. Korean J Spine. 2014 Sep;11(3):205-8. doi: 10.14245/kjs.2014.11.3.205.
75. Ahmed S, Paul SP. Intramedullary spinal neurocysticercosis treated successfully with medical therapy. J Egypt Soc Parasitol. 2014 Dec;44(3):661-4. doi: 10.12816/0007869.
76. Abarrategui Yagüe B, García García ME, Orviz García A, Casas Limón J. Lymphocytic meningitis and spinal neurocysticercosis: a case report and literature review. Neurologia. 2014 Nov-Dec;29(9):574-6. English, Spanish. doi: 10.1016/j.nrl.2013.02.005.
77. Iacoangeli M, Moriconi E, Gladi M, Scerrati M. Isolated cysticercosis of the cauda equina. J Neurosci Rural Pract. 2013 Aug;4(Suppl 1):S117-9. doi: 10.4103/0976-3147.116440.
78. Furtado SV, Dadlani R, Ghosal N, Rao AS. Solitary thoracic vertebral body cysticercosis presenting with progressive compressive myelopathy. J Neurosurg Spine. 2013 Apr;18(4):394-7. doi: 10.3171/2013.1.SPINE12675.
79. De Feo D, Colombo B, Dalla Libera D, Martinelli V, Comi G. Subarachnoid neurocysticercosis with spinal involvement presented with headache. Neurol Sci. 2013 Aug;34(8):1467-9. doi: 10.1007/s10072-012-1219-2.
80. Chandramohan R, Kadhiravan T, Swaminathan RP. Cysticercosis of the spinal cord. QJM. 2013 Mar;106(3):279-80. doi: 10.1093/qjmed/hcs010.
81. Araujo ABS, Cambraia MBR, Motta Filho RAM, Rezende GL, Vanderlei AS. Spinal intramedullary cysticercosis: a case report and literature review. Arquivos Brasileiros de Neurocirurgia: Brazilian Neurosurgery. 2013;32(04):245-9.
82. Shin SH, Hwang BW, Lee SJ, Lee SH. Primary extensive spinal subarachnoid cysticercosis. Spine (Phila Pa 1976). 2012 Sep 1;37(19):E1221-4. doi: 10.1097/BRS.0b013e31825d291e.
83. Rice B, Perera P. Intramedullary spinal neurocysticercosis presenting as brown-sequard syndrome. Western Journal of Emergency Medicine. 2012;13(5):434. DOI: 10.5811/westjem.2011.10.6909.
84. Naguib MM, Abramowsky CR, Shehata BM. Spinal cysticercosis mimicking a tumor in a pediatric patient. Fetal Pediatr Pathol. 2012 Apr;31(2):50-3. doi: 10.3109/15513815.2011.648724.
85. Motsepe T, Ackerman D. Spinal and vertebral neurocysticercosis in an HIV-positive female patient. Southern African Journal of Epidemiology and Infection. 2012;27(3):133-6.
86. Kapu R, Singh MK, Pande A, Vasudevan MC, Ramamurthi R. Intradural extramedullary cysticercal abscess of spine. Trop Parasitol. 2012 Jul;2(2):131-4. doi: 10.4103/2229-5070.105181.
87. Jain N, Gutch M, Agrawal A, Khanna A. Quadriparalytic disseminated neurocysticercosis. BMJ Case Rep. 2012 Jul 11;2012:bcr0820114613. doi: 10.1136/bcr.08.2011.4613.
88. Bhowmik A, Mondal B, Pal R, Banerjee P. A case of extensive neurocysticercosis involving brain and spinal cord. Journal of Pediatric Neurology. 2012;10(03):229-31. DOI 10.3233/JPN-2012-0560.
89. Agale SV, Bhavsar S, Choudhury B, Manohar V. Isolated intramedullary spinal cord cysticercosis. Asian J Neurosurg. 2012 Apr;7(2):90-2. doi: 10.4103/1793-5482.98655.
90. Vij M, Jaiswal S, Jaiswal AK, Behari S. Coexisting intramedullary schwannoma with intramedullary cysticercus: report of an unusual collision. Indian J Pathol Microbiol. 2011 Oct-Dec;54(4):866-7. doi: 10.4103/0377-4929.91541.
91. Seo JH, Seo HJ, Kim SW, Shin H. Isolated spinal neurocysticercosis : unusual ocular presentation mimicking pseudotumor cerebri. J Korean Neurosurg Soc. 2011 May;49(5):296-8. doi: 10.3340/jkns.2011.49.5.296.
92. Qi B, Ge P, Yang H, Bi C, Li Y. Spinal intramedullary cysticercosis: a case report and literature review. International journal of medical sciences. 2011;8(5):420.
93. Park YS, Lee JK, Kim JH, Park KC. Cysticercosis of lumbar spine, mimicking spinal subarachnoid tumor. Spine J. 2011 Apr;11(4):e1-5. doi: 10.1016/j.spinee.2011.02.009.
94. Lambertucci JR, Vale TC, Pereira AC, Sousa-Pereira SR, Dias JC, Pedrosa MS, Oliveira MM. Teaching NeuroImages: isolated cervical spinal cord cysticercosis. Neurology. 2011 Dec 6;77(23):e138. doi: 10.1212/WNL.0b013e31823b4753.
95. Jongwutiwes U, Yanagida T, Ito A, Kline SE. Isolated intradural-extramedullary spinal cysticercosis: a case report. J Travel Med. 2011 Jul-Aug;18(4):284-7. doi: 10.1111/j.1708-8305.2011.00535.x.
96. Heredia Moy K, Oviedo Gamboa I, Panozo Borda SV, Zegarra Santiesteban W, Ricaldez Muñoz R, Villarroel Arze T. Spinal neurocysticercosis: imaging diagnosis, report of a clinical case. Gaceta Médica Boliviana. 2011;34(2):96-8.
97. Azfar SF, Kirmani S, Badar F, Ahmad I. Isolated intramedullary spinal cysticercosis in a 10-year-old female showing dramatic response with albendazole. J Pediatr Neurosci. 2011 Jan;6(1):52-4. doi: 10.4103/1817-1745.84409.
98. Ahuja B, Banerjee AK, Kak VK. Cysticercosis of filum terminale. Neurol India. 2011 Nov-Dec;59(6):922-3. doi: 10.4103/0028-3886.91388.
99. Lin J, Chu W, Ye X. Use of suction to treat intramedullary spinal cysticercosis. BMJ Case Rep. 2010;2010:bcr04.2009.1755. doi: 10.1136/bcr.04.2009.1755.
100. Lim BC, Lee RS, Lim JS, Cho KY. A case of neurocysticercosis in entire spinal level. J Korean Neurosurg Soc. 2010 Oct;48(4):371-4. doi: 10.3340/jkns.2010.48.4.371.
101. Kumar S, Handa A, Chavda S, Tiwari R, Abbey P. Intramedullary cysticercosis. J Clin Neurosci. 2010 Apr;17(4):522-3. doi: 10.1016/j.jocn.2009.04.026.
102. Jang JW, Lee JK, Lee JH, Seo BR, Kim SH. Recurrent primary spinal subarachnoid neurocysticercosis. Spine (Phila Pa 1976). 2010 Mar 1;35(5):E172-5. doi: 10.1097/BRS.0b013e3181b9d8b6.
103. Gonçalves FG, Neves PO, Jovem CL, Caetano C, Maia LB. Chronic myelopathy associated to intramedullary cysticercosis. Spine (Phila Pa 1976). 2010 Mar 1;35(5):E159-62. doi: 10.1097/BRS.0b013e3181c89f2c.
104. Dhillon RS, McKelvie PA, Wang YY, Han T, Murphy M. Cystic lesion of the ventriculus terminalis in an adult. J Clin Neurosci. 2010 Dec;17(12):1601-3. doi: 10.1016/j.jocn.2010.04.026.
105. Choi KB, Hwang BW, Choi WG, Lee SH. Herniated lumbar disc combined with spinal intradural extramedullary cysticercosis. J Korean Neurosurg Soc. 2010 Dec;48(6):547-50. doi: 10.3340/jkns.2010.48.6.547.
106. Shin DA, Shin HC. A case of extensive spinal cysticercosis involving the whole spinal canal in a patient with a history of cerebral cysticercosis. Yonsei Med J. 2009 Aug 31;50(4):582-4. doi: 10.3349/ymj.2009.50.4.582.
107. Gupta S, Singh PK, Gupta B, Singh V, Azam A. Isolated primary intradural extramedullary spinal neurocysticercosis: a case report and review of literature. Acta Neurol Taiwan. 2009 Sep;18(3):187-92.
108. Chhiber SS, Singh B, Bansal P, Pandita KK, Razdan S, Singh J. Intramedullary spinal cysticercosis cured with medical therapy: case report and review of literature. Surg Neurol. 2009 Dec;72(6):765-8; discussion 768-9. doi: 10.1016/j.surneu.2009.06.011.
109. Mohapatra RN, Pattanaik JK, Satpathy SK, Joshi S. Isolated and silent spinal neurocysticercosis associated with pseudotumor cerebri. Indian J Ophthalmol. 2008 May-Jun;56(3):249-51. doi: 10.4103/0301-4738.40372.
110. Kasliwal MK, Gupta DK, Suri V, Sharma BS, Garg A. Isolated spinal neurocysticercosis with clinical pleomorphism. Turkish Neurosurgery. 2008;18(3):294-7.
111. Izci Y, Moftakhar R, Salamat MS, Baskaya MK. Spinal intramedullary cysticercosis of the conus medullaris. Wisconsin Medical Journal (WMJ). 2008;107(1):37.
112. Edwards CM, Miranda A, Pezzo S. Intradural-extramedullary spinal cysticercosis without parenchymal involvement. American Journal of Case Reports. 2008;9:301-3. http://www.amjcaserep.com/fulltxt.php?ICID=863371.
113. Agrawal R, Chauhan SPS, Misra V, Singh PA, Gopal NN. Focal spinal intramedullary cysticercosis. ACTA BIOMEDICA-ATENEO PARMENSE. 2008;79(1):39.
114. Paterakis KN, Kapsalaki E, Hadjigeorgiou GM, Barbanis S, Fezoulidis I, Kourtopoulos H. Primary spinal intradural extramedullary cysticercosis. Surg Neurol. 2007 Sep;68(3):309-11; discussion 312. doi: 10.1016/j.surneu.2006.10.060.
115. Ahmad FU, Sharma BS. Treatment of intramedullary spinal cysticercosis: report of 2 cases and review of literature. Surg Neurol. 2007 Jan;67(1):74-7; discussion 77. doi: 10.1016/j.surneu.2006.03.034.
116. Rossi LA, Sestari A, Cerioni Jr M. Intradural-extramedullary spinal cysticercosis with brain involvement: A case report and literature review. Radiologia Brasileira. 2006;39(5):379-82. DOI: 10.1590/s0100-39842006000500015.
117. Guedes-Corrêa JF, Macedo RC, Vaitsman RP, Mattos JG, Agra JM. Intramedullary spinal cysticercosis simulating a conus medullaris tumor: case report. Arq Neuropsiquiatr. 2006 Mar;64(1):149-52. doi: 10.1590/s0004-282x2006000100033.
118. Kim S-W, Lee S-M. Sacral intradural cysticercosis misdiagnosed as brain tumor metastasis. Journal of Korean Neurosurgical Society. 2005;37(1):67-9.
119. Torabi AM, Quiceno M, Mendelsohn DB, Powell CM. Multilevel intramedullary spinal neurocysticercosis with eosinophilic meningitis. Arch Neurol. 2004 May;61(5):770-2. doi: 10.1001/archneur.61.5.770.
120. Singh P, Sahai K. Intramedullary cysticercosis. Neurol India. 2004 Jun;52(2):264-5.
121. Jarupant W, Sithinamsuwan P, Udommongkol C, Reuarrom K, Nidhinandana S, Suwantamee J. Spinal cord compression and bilateral sensory neural hearing loss: an unusual manifestation of neurocysticercosis. JOURNAL-MEDICAL ASSOCIATION OF THAILAND. 2004;87(10):1244-9. http://www.medassocthai.org/journal
122. Delobel P, Signate A, El Guedj M, Couppie P, Gueye M, Smadja D, Pradinaud R. Unusual form of neurocysticercosis associated with HIV infection. Eur J Neurol. 2004 Jan;11(1):55-8. doi: 10.1046/j.1351-5101.2003.00696.x.
123. Jang HD, Park KH, Kim JC. Cervical Intramedullary Cysticercosis: Case Report. Journal of Korean Neurosurgical Society. 2003;33(3):323-5.
124. Costa Junior LBd, Lemos SP, Lambertucci JR. Magnetic resonance imaging of racemous cysticercosis of the cauda equina. Revista da Sociedade Brasileira de Medicina Tropical. 2003;36:765-6.
125. Sheehan JP, Sheehan J, Lopes MB, Jane JA Sr. Intramedullary spinal cysticercosis. Case report and review of the literature. Neurosurg Focus. 2002 Jun 15;12(6):e10. doi: 10.3171/foc.2002.12.6.11.
126. Yoon JW, Rhee DY, Park H, Park HS, Kim SY. A Case of Neurocysticercosis Presenting with Myelopathy and Hydrocephalus. Journal of Korean Neurosurgical Society. 2002;31(1):82-5.
127. Muzumdar D, Nadkarni T, Desai K, Dindorkar K, Goel A. Thoracic intramedullary cysticercosis: Two case reports. Neurologia Medico-Chirurgica. 2002;42(12):575-9. DOI: 10.2176/nmc.42.575.
128. Alsina GA, Johnson JP, McBride DQ, Rhoten PR, Mehringer CM, Stokes JK. Spinal neurocysticercosis. Neurosurg Focus. 2002 Jun 15;12(6):e8. doi: 10.3171/foc.2002.12.6.9.
129. Parmar H, Shah J, Patwardhan V, Patankar T, Patkar D, Muzumdar D, Prasad S, Castillo M. MR imaging in intramedullary cysticercosis. Neuroradiology. 2001 Nov;43(11):961-7. doi: 10.1007/s002340100615.
130. Mathuriya SN, Khosla VK, Vasishta RK, Tewari MK, Pathak A, Prabhakar S. Intramedullary cysticercosis : MRI diagnosis. Neurol India. 2001 Mar;49(1):71-4.
131. Homans J, Khoo L, Chen T, Commins DL, Ahmed J, Kovacs A. Spinal intramedullary cysticercosis in a five-year-old child: case report and review of the literature. Pediatr Infect Dis J. 2001 Sep;20(9):904-8. doi: 10.1097/00006454-200109000-00016..
132. Sahoo PK. Spinal intramedullary cysticercosis. Med J Armed Forces India. 2000 Jul;56(3):240-241. doi: 10.1016/S0377-1237(17)30178-8.
133. Gaur V, Gupta RK, Dev R, Kathuria MK, Husain M. MR imaging of intramedullary spinal cysticercosis: A report of two cases. Clin Radiol. 2000 Apr;55(4):311-4. doi: 10.1053/crad.1999.0080.
134. Dantas FL, Fagundes-Pereyra WJ, De Souza CT, Vega MG, De Souza AA. Intramedullary cysticercosis: case report. Arquivos de neuro-psiquiatria. 1999;57(2 A):301-5.
135. Ciftçi E, Diaz-Marchan PJ, Hayman LA. Intradural-extramedullary spinal cysticercosis: MR imaging findings. Comput Med Imaging Graph. 1999 May-Jun;23(3):161-4. doi: 10.1016/s0895-6111(99)00005-1.
136. Rosahl S, Samii M. Spinal intramedullary neurocysticercosis: Fallbericht. Klinische Neuroradiologie. 1998;8:115-22.
137. Mohanty A, Das S, Kolluri VR, Das BS. Spinal extradural cysticercosis: a case report. Spinal Cord. 1998 Apr;36(4):285-7. doi: 10.1038/sj.sc.3100524.
138. Lau KY, Roebuck DJ, Mok V, Ng HK, Lam J, Teo JG, Kay R, Poon W, Metreweli C. MRI demonstration of subarachnoid neurocysticercosis simulating metastatic disease. Neuroradiology. 1998 Nov;40(11):724-6. doi: 10.1007/s002340050672.
139. Garg RK, Nag D. Intramedullary spinal cysticercosis: response to albendazole: case reports and review of literature. Spinal Cord. 1998 Jan;36(1):67-70. doi: 10.1038/sj.sc.3100526.
140. Escobar A, Herrera MP, Escobar W, Vega R. Spinal intramedullary cysticercosis: a case report. Neuropathology. 1998;18(3):343-6.
141. Davies MA, Turner J, Bentivoglio P. Spinal and basilar extraparenchymal neurocysticercosis. J Clin Neurosci. 1996 Apr;3(2):174-7. doi: 10.1016/s0967-5868(96)90014-4.
142. Corral I, Quereda C, Moreno A, López-Vélez R, Martínez-San-Millán J, Guerrero A, Sotelo J. Intramedullary cysticercosis cured with drug treatment. A case report. Spine (Phila Pa 1976). 1996 Oct 1;21(19):2284-7. doi: 10.1097/00007632-199610010-00023.
143. Prasathapong S. Intramedullary cysticercosis: first case report in Thailand and literature review. Chulalongkorn Medical Journal. 1995;39(6):443-50. https://digital.car.chula.ac.th/clmjournal/vol39/iss6/6.
144. Kim SC, Han MH, Chang KH, Han GS, Hwang HY. MRI of intraspinal cysticercosis. Journal of the Korean Radiological Society. 1995;32(1):33-7.
145. Gallani NR, Zambelli HJ, Roth-Vargas AA, Limoli Júnior C. Cisticercose medular. Relato de dois casos, revisão da literatura e comentários sobre a patogenia [Spinal cord cysticercosis: report of 2 cases, review of the literature, and comments on its pathogeny]. Arq Neuropsiquiatr. 1992 Sep;50(3):343-50. Portuguese. doi: 10.1590/s0004-282x1992000300014.
146. Bandres JC, White AC Jr, Samo T, Murphy EC, Harris RL. Extraparenchymal neurocysticercosis: report of five cases and review of management. Clin Infect Dis. 1992 Nov;15(5):799-811. doi: 10.1093/clind/15.5.799.
147. Palasis S, Drevelengas A. Extramedullary spinal cysticercosis. Eur J Radiol. 1991 May-Jun;12(3):216-8. doi: 10.1016/0720-048x(91)90075-7.
148. Venkataramana NK, Jain VK, Das BS, Rao TV. Intramedullary cysticercosis. Clin Neurol Neurosurg. 1989;91(4):337-41. doi: 10.1016/0303-8467(89)90011-5.
149. Sperlescu A, Balbo RJ, Rossitti SL. Brief comments on the pathogenesis of spinal cysticercosis. Arquivos de neuro-psiquiatria. 1989;47(1):105-9.
150. Vlok G, Wells M. Vertebral cysticercosis-a case report. South African Medical Journal. 1988;73(12):730-1.
151. Castillo M, Quencer RM, Post MJ. MR of intramedullary spinal cysticercosis. AJNR Am J Neuroradiol. 1988 Mar-Apr;9(2):393-5.
152. Sharma BS, Banerjee AK, Kak VK. Intramedullary spinal cysticercosis. Case report and review of literature. Clin Neurol Neurosurg. 1987;89(2):111-6. doi: 10.1016/0303-8467(87)90185-5.
153. Savoiardo M, Cimino C, Passerini A, La Mantia L. Mobile myelographic filling defects: spinal cysticercosis. Neuroradiology. 1986;28(2):166-9. doi: 10.1007/BF00327891.
154. Holtzman RN, Hughes JE, Sachdev RK, Jarenwattananon A. Intramedullary cysticercosis. Surg Neurol. 1986 Aug;26(2):187-91. doi: 10.1016/0090-3019(86)90375-7.
155. Kim KS, Weinberg PE. Spinal cysticercosis. Surg Neurol. 1985 Jul;24(1):80-2. doi: 10.1016/0090-3019(85)90070-9.
156. McDonald JB, Turner PT, Miller AH. Cysticercosis and spinal cord compression. Ann Neurol. 1979 Oct;6(4):367-8. doi: 10.1002/ana.410060414.
157. Akiguchi I, Fujiwara T, Matsuyama H, Muranaka H, Kameyama M. Intramedullary spinal cysticercosis. Neurology. 1979 Nov;29(11):1531-4. doi: 10.1212/wnl.29.11.1531.
158. Firemark HM. Spinal cysticercosis. Arch Neurol. 1978 Apr;35(4):250-1. doi: 10.1001/archneur.1978.00500280068016.
159. Carmalt JE, Theis J, Goldstein E. Spinal cysticercosis. West J Med. 1975 Oct;123(4):311-3.
160. Singh A, Aggarwal ND, Malhotra KC, Puri DS. A case of spinal cysticercosis with paraplegia. Br Med J. 1966 Sep 17;2(5515):684-5. doi: 10.1136/bmj.2.5515.684.
161. Hesketh KT. Cysticercosis of the dorsal cord. J Neurol Neurosurg Psychiatry. 1965 Oct;28(5):445-8. doi: 10.1136/jnnp.28.5.445.
162. Cruz OR. Spinal cysticercosis: report of two cases with surgical treatment. Arquivos de Neuro-Psiquiatria. 1961;19:231-5.
163. Barini O. Intraspinal macrocystic cysticercus; surgical removal. Arquivos de neuro-psiquiatria. 1954;12(3):264-6.
